# Landmark ctDNA molecular response represents an early predictor of immunotherapy outcomes in lung cancer: A clinical utility study

**DOI:** 10.64898/2026.02.18.26346415

**Authors:** Noushin Niknafs, Lavanya Sivapalan, Archana Balan, Jaime Wehr, Gavin Pereira, Samira Hosseini-Nami, Nisha Rao, Shreshtha Jolly, Kavya Velliangiri, Iiasha Beadles, Tiffany Loftus, Bryan Chesnick, Jamie Medina, Wenming Xiao, Aliyah Pabani, Kristen A. Marrone, Qing Kay Li, Joseph C. Murray, Lorenzo Rinaldi, Nicholas C. Dracopoli, Mark Sausen, Christine L. Hann, Susan C. Scott, Josephine Feliciano, Vincent K. Lam, Benjamin Levy, Victor E. Velculescu, Julie R. Brahmer, Patrick M. Forde, Paz J. Vellanki, Valsamo Anagnostou

## Abstract

**Purpose:** Circulating tumor DNA (ctDNA) analyses are informative as an early indicator of immunotherapy response in advanced non-small cell lung cancer (NSCLC); however, the clinical value of ctDNA molecular response requires further validation.

**Patients and Methods:** As part of a prospective clinical protocol (NCT05995821), we conducted targeted error-correction sequencing of ctDNA (n=328) and matched WBC DNA (n=109) from 109 patients with metastatic NSCLC who received anti-PD-(L)1 either as monotherapy or in combination. Following cellular origin resolution of 2,818 variants, landmark molecular response (mR) was defined as undetectable ctDNA within 3-9 weeks of treatment initiation.

**Results:** Pre-treatment ctDNA burden, but not blood tumor mutation burden, predicted survival. Implementing a tumor-naïve WBC DNA-informed approach increased the number of evaluable cases without compromising the overall accuracy of landmark ctDNA molecular responses. A direct comparison of single-timepoint on-therapy ctDNA assessment with ctDNA dynamics from baseline to the 3-9-week interval, along with an analysis of heterogeneity in molecular response within the 3-9-week window, showed that undetectable ctDNA at the landmark timepoint can effectively predict survival outcomes. A significant enrichment in landmark ctDNA mR was noted among patients with progression-free survival (PFS) ≥6 months with immunotherapy (p=2.5e-05) and chemo-immunotherapy (p=0.02). Patients in the landmark mR group had longer progression-free (p=1.6e-06) and overall survival (p=2.5e-05) than those with molecular progression.

**Conclusions:** Landmark ctDNA molecular response provides a real-time, accurate approach for monitoring immunotherapy clinical outcomes. Although not currently validated for regulatory use, these findings demonstrate the potential utility of ctDNA as an early endpoint in clinical trials.

**Translational Relevance:** Employing circulating tumor DNA (ctDNA) dynamics as an early indicator of immunotherapy response requires a roadmap for the next-generation sequencing approach, definition of molecular response and establishment of its clinical sensitivity. In this study, we introduce the concept of a landmark ctDNA molecular response, determined 3-9 weeks after initiation of immunotherapy, that maximizes the number of evaluable patients without sacrificing the specificity of the approach. Notably, when evaluating heterogeneity in ctDNA detection within the landmark 3-9-week window and assessing the impact of landmark interval dynamics on survival, we found that a single ctDNA assessment performed similarly to multiple ctDNA measurements within the landmark window (most notably, regardless of whether the timepoints were concordant or discordant). Our findings demonstrate that a single assessment of early on-therapy landmark ctDNA molecular response, can identify patients at risk of disease progression and enable future intervention and therapy optimization.

## Introduction

The landscape of immuno-oncology (IO) is rapidly evolving, exemplified by breakthroughs in the treatment of patients with both earlier-stage and advanced non-small-cell lung cancer (NSCLC) (1–7). These advancements underscore the critical importance of ongoing evaluation, utilization, and refinement of robust biomarkers to accurately monitor therapy responses and guide treatment decisions (8, 9). However, assessing therapy outcomes in the context of immunotherapy presents unique challenges, including the heterogeneity of clinical responses and the nuanced nature of tumor kinetics that are inadequately captured by conventional imaging (8, 9). Early changes in circulating tumor DNA (ctDNA), particularly ctDNA clearance, have been shown to serve as a reliable early predictor of response to immune checkpoint inhibitors (ICI) (10–13). We have shown the clinical value of ctDNA clearance after 6 weeks of pembrolizumab monotherapy, as an early predictor of immunotherapy response in the context of a ctDNA-adaptive clinical trial for patients with treatment-naïve driver oncogene-negative NSCLC (13). Timely detection of disease progression on immunotherapy is pivotal, as it provides a therapeutic window during which patients with primary resistance can be promptly identified and redirected towards alternative treatment strategies (8, 13).

Yet, several key considerations persist, such as determining the cellular origin of mutations in cell-free DNA (cfDNA), identifying the optimal metric for monitoring cell-free tumor load, assessing the effectiveness of tumor-agnostic versus tumor-informed approaches, and studying the ctDNA kinetics by histological subtypes of NSCLC or treatment modalities (14). Importantly, the definition of ctDNA molecular response varies by study cohort and assay used, and historically, ctDNA molecular response has relied on analyses of two timepoints: baseline and early on-therapy (11). Considering the potential cost-effectiveness of serial liquid biopsies compared to the cost of ICI therapy and radiographic imaging, early determination of ctDNA response based on analyses of the most informative timepoint could offer significant benefits to patients and healthcare systems. Furthermore, while use of repeat plasma comprehensive genomic profiling is established in the context of oncogene-driven NSCLC (15), the clinical utility of liquid biopsies in detecting emerging mechanisms of immunotherapy resistance is not well established.

In addition to testing ctDNA molecular response in ctDNA-adaptive clinical trials, a critical step towards validating liquid biopsy approaches and their practical implementation in real-world settings is to leverage prospective biobanking protocols. The Johns Hopkins Immunobiology Blood and Tissue Collection protocol (NCT05995821) was designed to prospectively enrol patients with thoracic cancers, harmonize the collection of serial blood biospecimens and streamline minimally invasive studies across histologies and treatment modalities. As part of these efforts, we evaluated the evolving ctDNA landscape during therapy and prospectively tested the clinical sensitivity of the landmark ctDNA molecular response in predicting immunotherapy outcomes.

## Patients and Methods

### Clinical protocol and cohort characteristics

A cohort of 109 patients, who were prospectively enrolled in the Immunobiology Blood and Tissue Collection of Upper Aerodigestive Malignancies clinical protocol (NCT05995821) between October 2016 and April 2022, were included in the study (table S1). Key eligibility criteria included (1) confirmed histological diagnosis of NSCLC, (2) locally advanced or metastatic disease, (3) ICI therapy at the time of enrolment, (4) availability of at least 2 serial plasma samples or a single sample within the 3-9-week window for cell-free DNA (cfDNA) sequencing, and (5) presence of at least one matched buffy-coat sample for white blood cell (WBC) genomic DNA sequencing. Blood samples were collected at predefined intervals, approximately every 3-6 weeks, until disease progression (n=42), treatment completion (n=5), discontinuation due to toxicity (n=29), loss to follow-up (n=2), treatment break/surveillance (n=14), or initiation of maintenance immunotherapy (n=17). Clinical data were retrieved from electronic health records through the Johns Hopkins Lung Cancer Precision Medicine Center of Excellence. A total of 83 patients were therapy-naïve before receiving ICI therapy, while 26 patients received ICI-containing regimens as second-line or later treatments. One patient harboured a canonical activating exon 19 EGFR mutation and received ICI therapy after disease progression on EGFR targeted therapy. Progression-free survival (PFS) and overall survival (OS) were evaluated relative to the initiation time for immune checkpoint inhibitor treatment, along with landmark PFS determination at 6 months as the clinical study endpoints. Patients with a PFS ≥6 moths were classified in the durable clinical benefit (DCB) group, while patients with PFS <6 months in the non-durable clinical benefit (NDB) group. Six-month landmark PFS could not be evaluated in four patients who had a follow-up period of less than 6 months, and these patients were excluded from correlative assessments. This study was conducted in accordance with the Declaration of Helsinki, approved by the Institutional Review Board (IRB), and patients provided written informed consent for the collection of samples, clinical data and liquid biopsy analyses for research purposes. The clinicopathological characteristics of all study participants are summarized in Supplementary Table S1.

### Blood sample processing, cfDNA and WBC DNA sequencing

Blood samples collected during routine venepuncture in K2EDTA tubes underwent processing for plasma and buffy-coat isolation, followed by targeted error-correction sequencing (TEC-Seq) (10) (tables S2-S4). In brief, plasma cfDNA was extracted from 0.8-4.8 ml plasma using the MagMAX™ Cell-Free DNA Isolation kit (Life Technologies; Austin, TX, USA). Matched WBC DNA was isolated from buffy-coat samples of all patients, using the Qiagen DNA Blood Mini kit (Qiagen GmbH), and then sheared to a target fragment size of 200 bp. TEC-Seq DNA libraries were generated from 2.6 - 26 ng of cfDNA or WBC DNA (table S2), followed by targeted capture using a customized set of hybridization probes (Elio Plasma Complete, Personal Genome Diagnostics, Baltimore, MD; table S3) according to the manufacturer’s protocol. Libraries were pooled and sequenced with 150 bp paired-end reads using the Illumina NovaSeq6000 platform attaining 2,500-fold de-duplicated coverage across the target regions. Samples with 7-26 ng of input DNA from individual patients were batched together for processing, while those with <7 ng input DNA were processed separately. Two patients had insufficient cfDNA input. TEC-Seq characteristics for plasma and matched WBC DNA samples are summarized in table S4.

### Whole exome sequencing

Whole exome sequencing (WES) of matched tumor tissue was performed for 34 patients with sufficient tumor tissue as previously described (16, 17). Tumor DNA extraction followed pathological review and macro-dissection of formalin-fixed paraffin-embedded (FFPE) tumor tissue, using the Qiagen DNA FFPE kit (Qiagen GmbH). Tumor and matched WBC DNA were sheared to a target fragment size of 200 bp and processed for Illumina TruSeq library construction (Illumina). Hybrid capture of exonic regions was performed using Agilent SureSelect in-solution capture reagents and SureSelect XT Human All Exon V4 probes (Agilent). Captured libraries underwent sequencing with 100 bp paired-end runs on Illumina HiSeq 2500 instruments. Somatic mutations were identified by paired tumor-normal analysis following an approach described previously (16, 17). Importantly, a minimum tumor mutant allele fraction of 10% in the tumor sample was required to filter out sequencing and alignment artifacts. To perform tumor-informed plasma mutation analyses, plasma mutations at each timepoint were narrowed down to those identified in patient’s matched tumor sample.

### Analysis of plasma sequencing data and classification of somatic variant cellular origin

TEC-Seq data were analysed using the PGDx elio platform software (Personal Genome Diagnostics, Baltimore, MD). Somatic variant identification was performed following error-correction as previously described (16, 18). The variant allele fraction (VAF) for a given plasma variant was determined by calculating the percentage of distinct mutant versus total cfDNA variant reads at a given locus. The origin of plasma cfDNA variants was classified in a tumor-naive WBC DNA-informed manner, as tumor-derived, germline, or clonal haematopoiesis (CH)-derived (fig. s1). To characterize co-occurring genomic alterations in ctDNA, tumor-derived mutations in plasma were aggregated to the gene level and compared with frequently co-occurring genomic alterations in lung cancer (19). Mutations were selected for blood tumor mutation burden (bTMB) measurement based on the following assay-specific criteria: somatic nonsynonymous single nucleotide variants (SNVs) with a variant allele frequency of at least 1% and a minimum supermutant read count of 6. Mutations located in COSMIC hotspot regions, putative CH variants, and common germline polymorphisms (reported in dbSNP or gnomAD) were excluded to ensure the specificity of bTMB assessment. Of note, baseline bTMB could not be computed for two patients (CGLU295 and CGLU329), but baseline maxMAF was available.

### Assessment of cfDNA fragment lengths

To further improve the accuracy of variant origin classification, we utilized fragment length distributions containing mutant alleles compared to wild-type cfDNA fragments and built a logistic regression model for variant cellular origin prediction (fig. S2A). Briefly, we used the subset of patients with tumor whole-exome sequencing data to compile a training set comprising 458 validated tumor-derived somatic mutations. Furthermore, we identified 5772 common SNPs (dbSNP database, build ID 151) in the matched WBC data with mutant allele fraction in 50-70% range expected for germline variants which passed all sequencing quality metrics. The training set comprised of these 458 validated tumor-derived variants and 5772 high confidence germline variants. For each mutation, we extracted the fragments harbouring the mutant or wild-type allele in plasma cfDNA, and calculated the ratio between the median of mutant fragment lengths and the median of wild-type fragment lengths. To get an accurate estimate of the model’s generalizeability, we implemented a nested LOO cross-validation training setup. In each iteration, all plasma variants from a given patient were set aside (outer loop), and the parameters of the logistic regression model were estimated using a 10×5 training setup (10 repeats of 5 fold CV) in the inner loop. The final model resulting from the inner loops training was used to generate the probability scores for the hold-out plasma variants (outer loop), offering an unbiased estimate of the model’s performance in previously unseen variants (fig. S2B-C, table S6). For the variants that were not part of the training set (n=2304), we used the locked model generated by training on the entire training set to predict the probability of being tumor-derived for each variant (fig. S2D-E, table S7).

### Assessment of somatic copy number aberrations and plasma aneuploidy

Genome-wide copy number profiles of each cfDNA sample were analyzed as previously described (16). Briefly, sequencing reads mapped to target regions and background reads were used to construct a coverage depth map, corrected for GC content, region size, and sequence mappability. Somatic copy number profiles were evaluated using matched WBC DNA samples (n=109) to establish a reference panel. The relative copy number profile for each genomic region was derived and Circular Binary Segmentation was applied to identify segments with constant copy numbers. Plasma aneuploidy (PA) scores were calculated by adjusting the log copy ratio of segments to a mean of 0, transforming them into a linear scale, and computing weighted averages for each chromosomal arm. These values were then transformed into z-scores and combined to calculate the PA score per sample. Plasma aneuploidy (PA) scores from WBC DNA samples ranged from -1.91 to 2.97; therefore, a conservative PA threshold of 2.97 was selected to distinguish between normal ploidy and cancer aneuploidy in plasma. More specifically, PA scores were dichotomized as either indicative of plasma cancer aneuploidy (PA > 2.97) or normal ploidy (PA score ≤ 2.97).

### Longitudinal tracking of circulating tumor fraction and molecular response classification

Landmark molecular responses were calculated using the mutant allele fraction of the most abundant tumor-derived mutation (maxMAF) at the 3–9-week window following treatment initiation. We used a pre-determined landmark molecular response definition, denoted as the absence of detectable maxMAF (i.e. a maxMAF value of 0%, no detectable tumor-derived alterations) at any time point within 3-9 weeks from treatment initiation, irrespective of any fluctuations or changes in detectability at later timepoints within the landmark interval. Fifteen patients had plasma collections falling outside the 3-9-week window, rendering them not evaluable for landmark molecular response. Four additional patients were excluded from molecular response analysis due to samples failing QC criteria (insufficient cfDNA yield, n = 2), or the timing of sample collection (plasma samples available only at baseline and at the time of disease progression, n =2). Thirteen patients who received durvalumab as maintenance therapy for stage III NSCLC following definitive chemoradiotherapy were excluded from landmark molecular response analyses. Overall, 77 patients with detectable tumor alterations were included in molecular response assessments. Clinical sensitivity is defined as the fraction of patients with durable clinical benefit that have concordance molecular response assignment, and clinical specificity indicates the fraction of patients with non-durable clinical benefit that have a concordant molecular progressive disease assignment.

### Statistical analyses

Survival analyses were conducted using the Kaplan-Meier method to derive median point estimates, with survival curves compared using logrank testing. The impact of molecular responses on OS and PFS was assessed through univariate and multivariate Cox proportional hazards regression analyses. Categorical comparisons were performed using Fisher’s exact test, while non-parametric comparisons utilized the Mann-Whitney U test. The association between the derived baseline bTMB values and clinicopathological variables including molecular response, histology, age, and sex was evaluated. To capture the dose-dependent impact of TMB on survival, a set of quantile-based thresholds ranging from 20% to 90% were considered. At each threshold, the patients were categorized into bTMB-high and bTMB-low groups based on the baseline sample. At each threshold, a univariate Cox proportional hazards regression model was used to assess the association between bTMB category and progression-free or overall survival.Statistical significance was determined at two-sided p < 0.05, unless otherwise specified. All statistical analyses were carried out using R v.4.3.

## Results

### Importance of elimination of biological noise from clonal haematopoiesis mutations

We performed targeted error-correction sequencing on serial plasma (n=328) and matched WBC DNA (n=109) from 109 patients with locally advanced or metastatic NSCLC receiving ICI-containing therapy (**Fig. 1**, tables S1-S3, and Methods). Sequence alterations in ctDNA were analyzed at baseline, longitudinally (every 3-6 weeks) during therapy, and at the time of clinical disease progression (table S4). To definitively resolve the origin of cfDNA variants, we incorporated a branched logic that leverages matched plasma and WBC DNA deep sequencing, together with consideration of gene and mutation frequencies in solid tumors and myelodysplasia as well as differential fragment length distributions (Methods, tables S5-S7 and fig. S1). As cfDNA fragmentation profiles can harbour epigenetic signatures reflective of cancer lineage, we assessed cfDNA fragment length distributions and observed that fragments carrying mutant alleles were significantly shorter compared to wild-type, while CH variants could not be distinguished from wild-type based on fragment length alone (fig. S2). Leveraging these observations, we trained a logistic regression model to predict mutation cellular origin based on fragment length differences between mutant and wild-type alleles on a training set comprising of 458 bona fide tumor-derived mutations (as confirmed by tumor tissue exome sequencing) and 5772 high-confidence germline variants (Methods, tables S6-S7 and fig. S2). Most plasma variants were concordantly classified (73% for tumor-derived, 93% for germline and 76% for CH-derived variants; table S7 and fig. S2) between the branched logic and logistic regression models, suggesting that mutant fragment length profiles may be leveraged to inform variant cellular origin. Through the branched logic, plasma variants were classified as tumor-derived, germline or clonal haematopoiesis (CH)-derived (table S5).

**Fig. 1.**
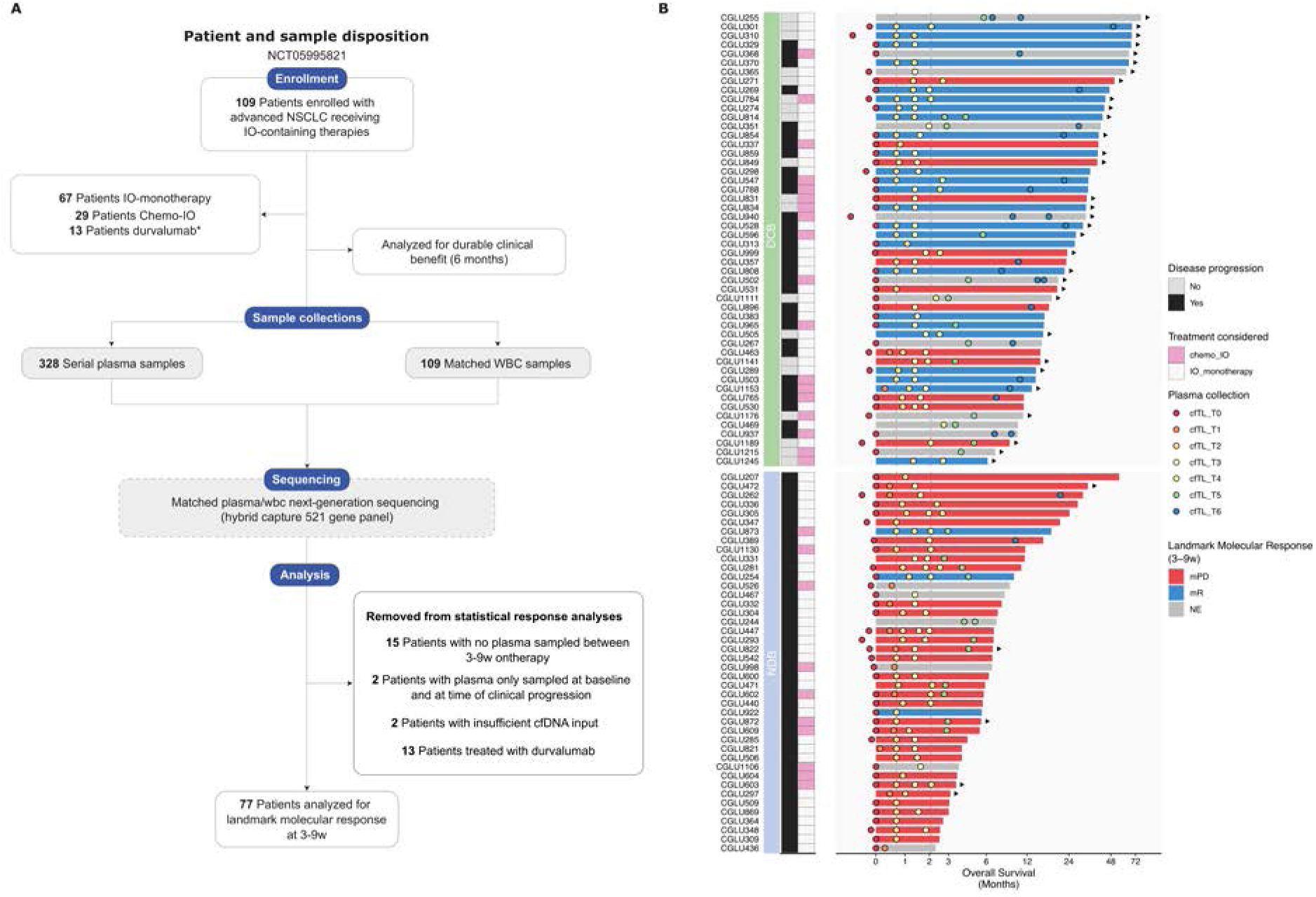
Patient enrolment and overview of study cohort. (**A**) Targeted error-correction sequencing was conducted on serial plasma (n=329) and matched white blood cell (WBC) genomic DNA samples (n=109) collected under the NCT05995821 protocol from 109 patients with advanced or metastatic NSCLC that received immunotherapy-containing treatments. Plasma variants were classified based on cellular origin (tumor-derived, germline, or clonal haematopoiesis (CH)-derived) using matched WBC DNA-informed filtering. Molecular response assessments were conducted within the 3–9-week window post-treatment initiation. Landmark molecular response (mR) was defined as absence of detectable ctDNA at any sampled timepoint within the 3-9 week interval, while persistent ctDNA marked molecular progressive disease (mPD). Two patients had insufficient cfDNA, thirteen had plasma collected outside the 3-9-week window, and two had only baseline and progression samples, rendering them unevaluable for molecular response assessment. Patients who received durvalumab as maintenance therapy following definitive chemoradiation for stage III disease were further removed from molecular response analyses. (**B**) Swimmer’s plot showing an overview of the evaluable study cohort according to landmark molecular response status and attaining PFS ≥6 months (marked as durable clinical benefit-DCB) or PFS <6 months (marked as non-durable clinical benefit-NDB). Plasma sampling timepoints are shown across relevant intervals assessed for the study (T0, pre-therapy; T1, >0–3 weeks; T2, 3-6 weeks; T3, 6-9 weeks; T4, 9-12 weeks; T5, 12-26 weeks; T6, >26 weeks). Four patients who could not be evaluated for clinical benefit due to follow-up < 6 months and patients treated with durvalumab are not shown. IO, immuno-oncology; mR, molecular response; mPD, molecular progressive disease; QC, quality control; cfTL: circulating tumor load.

We subsequently evaluated the impact of CH contamination on comprehensive genomic profiling of cfDNA and studied the landscape of cfDNA alterations in its entirety and by cellular origin. Of 2,818 plasma variants with resolved cellular origin, the majority, comprising 2,076 variants (74%), were classified as tumor-derived, 98 variants (3%) were identified as germline, and 644 variants (23%) were CH-derived (tables S7-S8). Genes frequently mutated in CH included *DNTM3A* (n=51 patients), *TP53* (n=14, **Fig. 2A**), *KMT2D* (n=5), *SMARCA4* (n=3, **Fig. 2B**), *MTOR* (n=3), *ERBB4* (n=3), and *ARID2* (n=3). Notably, of 148 alterations detected in *TP53*, 106 were deemed tumor-derived, while the remaining 42 variants were attributed to CH. The *TP53* CH-derived subset included 7 missense alterations (G245S, R158H, R175H, R248Q, R273H, R273L, Y220C) occurring at positions identical to tumor-derived *TP53* alterations identified across the cohort, demonstrating the necessity for matched WBC DNA-informed variant filtering to accurately distinguish such CH-variants from tumor-derived ctDNA mutations. Assessment of the prognostic value of baseline *TP53* and *SMARCA4* mutations with and without CH variant filtering revealed an association between *TP53* and *SMARCA4* tumor-derived mutations and poorer clinical outcomes, which was obliterated upon inclusion of CH mutations affecting these genes (**Fig. 2C-F).** These findings highlight the confounding role of CH variants, frequently detected by cfDNA comprehensive profiling and the importance of devising strategies for accurate classification of plasma variants by cellular origin.

**Fig. 2.**
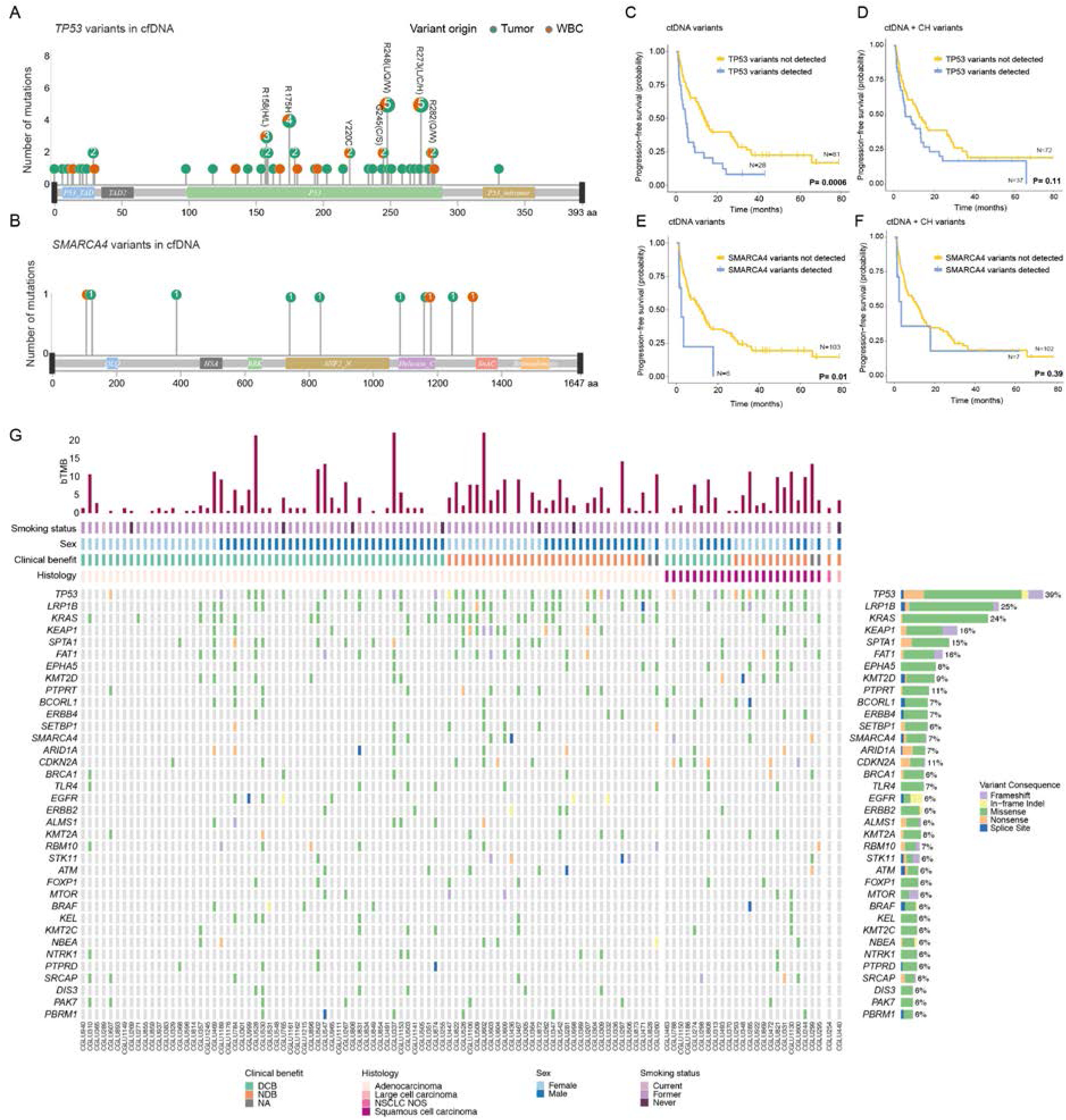
Landscape of sequence alterations detected in plasma cfDNA. (**A-B**) Positions and frequencies of tumor (*green*) and WBC (*orange*) variants detected across frequently mutated NSCLC driver genes (*TP53* and *SMARCA4*) demonstrate the prevalence of patient-specific CH alterations across hotspot loci, necessitating matched WBC DNA-informed variant filtering approaches combined with fragment length assessment to effectively filter out non-tumor CH variants from tumor-derived ctDNA. (**C-F**) Assessment of baseline plasma alterations in *TP53* and *SMARCA4* before and after CH filtering revealed that patients with ctDNA mutations in either driver gene had a significantly poorer progression-free survival (PFS) compared to those without detectable alterations. Importantly, these associations were diminished upon inclusion of CH alterations. (**G**) Overview of the landscape of ctDNA variants identified in plasma (>5% prevalence) according to histological subtype. Although *TP53* was the most frequently mutated gene across adenocarcinoma and squamous subtype tumors, the observed mutation rate in ctDNA was higher among cases with squamous cell carcinomas (52% vs 36%) compared to adenocarcinomas. In contrast, *KRAS* alterations were more frequent amongst patients with adenocarcinomas (29% vs 9%). Patients attaining PFS ≥6 months are marked as attaining durable clinical benefit (DCB), while patients with PFS <6 months as non-durable clinical benefit (NDB). cfDNA, cell-free DNA; ctDNA, CH, clonal haematopoiesis; circulating tumor DNA; mut, mutant; WBC, white blood cell; wt, wild-type.

### The compendium of ctDNA alterations captures the genomic landscape of NSCLC

Following resolution of variant origin and in analyses of tumor-derived mutations, 97 of 109 (89%) patients had ≥1 tumor-derived mutation detected at any sampled plasma timepoint, with a median mutant allele fraction of 2.2% (range 0.1% - 68.6%; table S5). The most frequent tumor-derived alterations were nonsynonymous mutations in *TP53* (39%), *LRP1B* (25%), *KRAS* (24%), *KEAP1* (16%) and *FAT1* (16%), consistent with the landscape of frequently mutated genes in non-oncogene addicted NSCLC (**Fig. 2G** and table S5). Alterations in *KRAS* and *TP53* frequently co-occurred with ctDNA mutations in *FAT1*, *KEAP1, SPTA1, PTPRT, PBRM1, MTOR, LRP1B, KMT2D, KMT2A, ERBB4* and *CDKN2A* (**Fig. 2G** and table S8). We then asked whether longitudinal plasma genomic profiling under the selective pressure of immunotherapy would reveal emerging genotypes that potentially confer therapy resistance. In 5 patients with ICI acquired resistance, defined as objective response or stable disease for at least 6 months followed by disease progression, emerging mutations were detected in *SMARCA4, KMT2C, BRCA1, RAD51, RAD51C* and *FOXP1* genes (fig. S3). These findings highlight the potential role of longitudinal plasma genotyping in capturing the emergence of cancer clones harbouring resistance mutations, such as in SMARCA4, or new putatively actionable genomic targets, which have been historically limited to oncogene-driven NSCLC.

### Molecular characterization of NSCLC histological subtypes by ctDNA comprehensive genomic profiling

We performed further analyses of ctDNA molecular landscapes to identify differences between adenocarcinomas (ACs) and squamous cell carcinomas (SCCs). In ACs, in addition to known driver mutations in *KRAS* (29%), recurrent mutations were identified in tumor suppressor genes, including *TP53* (36%), *KEAP1* (18%), *STK11* (6%), *FAT1* (15%), and *CDKN2A* (7%; **Fig. 2G**, fig. S4 and table S8). Recurring mutations in adenocarcinomas were further observed in genes involved in chromatin modification, including *ARID1A* (7%) and *SMARCA4* (6%), and the receptor tyrosine phosphatase *PTPRT* (13%; **Fig. 2G** and fig. S4). SCCs harboured a higher frequency of *TP53* mutations (52%) together with mutations in regulators of the oxidative stress response pathway [*KEAP1* (9%) and *NFE2L2* (9%)], mediators of squamous cell differentiation, including *FOXP1* (9%) and *NOTCH1* (13%), and chromatin regulating genes [*KMT2D* (17%), *KMT2A* (13%); **Fig. 2G**, fig. S4 and table S8]. *CDKN2A* mutations were more frequent in squamous cell carcinomas (26% vs 7% in adenocarcinomas), while *KRAS* ctDNA alterations and *KRAS* co-mutations were less prevalent among SCCs compared to ACs (9% vs 29% respectively; **Fig. 2G**, fig. S4 and table S8). In looking at ctDNA chromosomal arm-level somatic copy number alterations, patients with ACs and SCCs displayed gains of 1q, 7, 8q, and losses of 3p, 4p, 8p, 10q and 13q (fig. S4). Notably, a selective amplification of chromosome 3q characterized ctDNA from patients with SCCs (p = 0.008, Mann-Whitney U test; fig. S4 and table S9). In contrast, patients with adenocarcinomas displayed significant copy number losses across chromosomal arms 2p (Mann-Whitney p = 0.03), 2q (Mann-Whitney p = 0.02), and 20p (Mann-Whitney p = 0.01; fig. S4 and table S9).

### Baseline circulating tumor fraction but not bTMB is associated with immunotherapy outcomes

We subsequently employed the maximal mutant allele fraction of tumor-derived mutations (maxMAF) to estimate circulating tumor fraction at each timepoint studied. Assessment of baseline timepoints revealed a significant association between baseline maxMAF and survival outcomes, including clinical benefit, PFS, and OS (**Fig. 3A-C**, fig. S5 and table S5). A significant association was observed between baseline low maxMAF and clinical benefit (Fisher’s exact test p=0.005; **Fig. 3A**), which appeared to be driven by the subset of patients treated with first-line IO monotherapy (fig. S5B), while no associations were identified with histology, treatment, sex or race (fig. S5). Moreover, patients with a high baseline maxMAF (defined as ≥ the median baseline maxMAF of 2.59%) had shorter PFS (4.41 months vs 11.2 months, logrank p=0.011) and OS (8.22 months vs 33.01 months, logrank p=0.0035) compared to patients with a low baseline maxMAF (**Fig. 3B-C**). In contrast, we did not find analogous associations between baseline bTMB (coded as a continuous variable) and outcomes (Methods, **Fig. 3D** and table S10; all patients: hazard ratio-HR= 1.01, 95% confidence interval-CI 0.97-1.06, p=0.62 for PFS and HR=1.02, 95% CI 0.97-1.07, p=0.52 for OS; IO monotherapy subset: HR=1.00, 95% CI 0.94-1.07, p=0.97 for PFS and HR=0.99, 95% CI 0.92-1.07, p=0.84 for OS). Using a bTMB threshold of ≥16 mutations (20), we did not observe a significant association between high bTMB and OS (logrank p=0.85) or PFS (logrank p=0.97; fig. S5 and table S10). Similarly, using the median of baseline bTMB as the threshold did not show significant differences in PFS (HR= 0.8, 95% CI 0.48-1.35, p=0.40) or OS (HR=0.64, 95% CI 0.36-1.16, p=0.14) (fig. S6). These findings suggest that the circulating tumor load is predictive of immunotherapy outcomes, whereas the association between bTMB and therapy response remains equivocal.

**Fig. 3.**
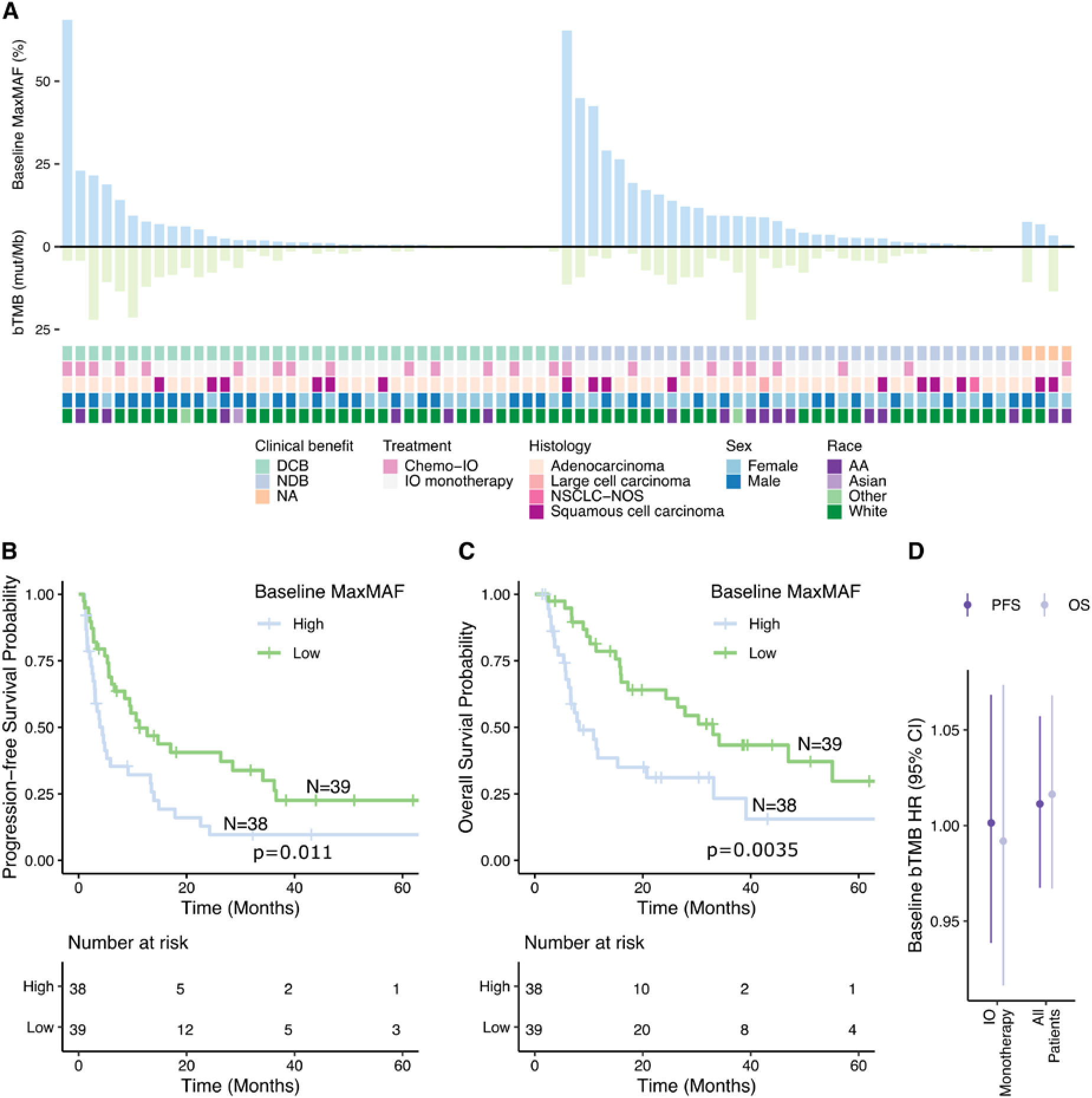
Baseline ctDNA characteristics are associated with clinical outcomes. (**A**) The maximum mutant allele fraction (maxMAF) of baseline ctDNA alterations and blood tumor mutation burden (bTMB) scores were evaluated in relation to clinicopathological variables and survival outcomes across the study cohort. Patients are sorted by clinical benefit status, and baseline maxMAF from left to right. Patients with PFS ≥6 months (marked as durable clinical benefit-DCB) had significantly lower baseline maxMAF, compared to patients with PFS <6 months (marked as non-durable clinical benefit-NDB, MW test p=0.005). Baseline maxMAF was also significantly elevated in patients with molecular progressive disease (mPD) compared to patients who attained a landmark molecular response (mR, p=0.01), whereas no significant differences were observed according to histology (p=0.4), sex (p=0.27) or race (p=0.66). (**B-C**) In patients with evaluable baseline plasma samples and detectable ctDNA (n=77), a high baseline maxMAF (> median 2.59%) was associated with a significantly poorer progression-free and overall survival compared to a low (<2.59%) baseline maxMAF (progression-free survival, logrank p=0.1, overall survival, logrank p=0.0035). (**D**) Univariate Cox proportional hazard analyses of progression-free and overall survival using baseline bTMB scores (coded as a continuous predictor) revealed no significant association (All patients, HR= 1.01, 95% CI 0.97-1.06, p=0.62 for PFS and HR=1.02, 95% CI 0.97-1.07, p=0.52 for OS; IO monotherapy subset, HR=1.00, 95% CI 0.94-1.07, p=0.97 for PFS and HR=0.99, 95% CI 0.92-1.07, p=0.84 for OS).

### Longitudinal dynamics of ctDNA sequence and structural landscapes reveal patterns of molecular response and progression

To assess longitudinal dynamic changes in circulating tumor fraction under the selective pressure of immunotherapy, we evaluated both sequence and structural alteration-based approaches (Methods). Dynamic changes in ctDNA levels computed by the tumor-agnostic WBC DNA-informed approach mirrored tumor-informed ctDNA dynamics (**Fig. 4A-B**, table S11, Methods) and identified clear patterns of ctDNA kinetics during therapy. For the assessment of plasma aneuploidy (PA), we examined chromosomal arm-level copy number profiles and utilized the most aberrant chromosomal arm representations in each sample to construct a PA score (Methods, table S12 and fig. S7). Plasma aneuploidy patterns were concordant with sequence alteration kinetics (fig. S7), such that PA shrunk to normal ploidy on-therapy for patients with maxMAF clearance (**Fig. 4C**), while remained elevated in cases with persistent detection of sequence alterations (**Fig. 4D-F**). Given the tight correlation between sequence and structural assessments of circulating tumor fraction (Pearson’s R=0.9, p=2.2e-16, fig. S7) and in line with our previous studies (13), we defined landmark molecular response (mR) as the absence of detectable maxMAF in at least one time point within 3-9 weeks from treatment initiation. Patients exhibiting ctDNA persistence within this interval were classified in the landmark molecular progression group (mPD). Overall, 77 patients had plasma sampled between 3-9 weeks on-therapy and were evaluable for molecular response assessment. Of these, 29 patients (38%) attained a mR, while 48 patients (62%) were classified as having mPD.

**Fig. 4.**
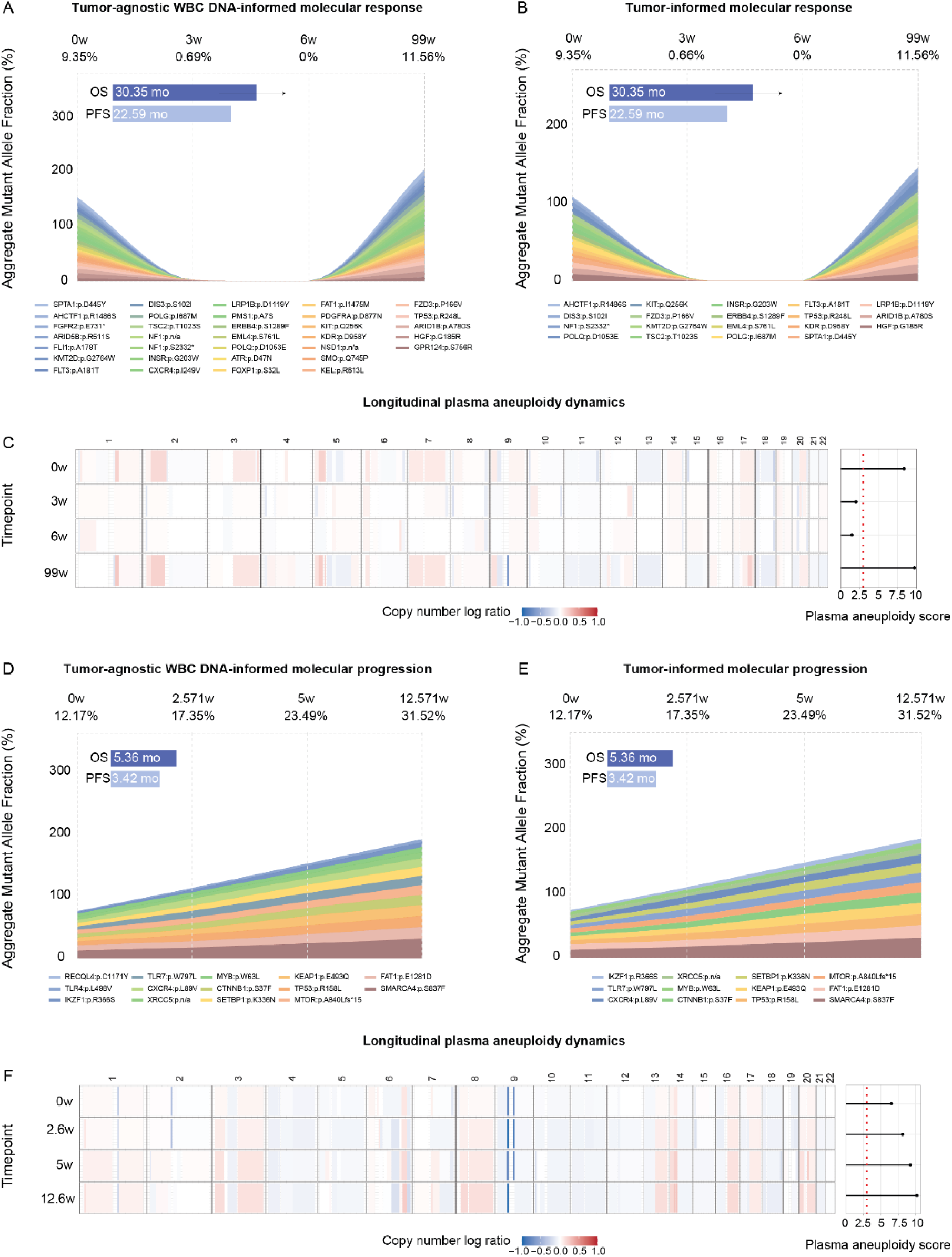
Longitudinal ctDNA dynamics capture early on-therapy molecular responses to immune checkpoint blockade. Dynamic changes in ctDNA were tracked across early on-therapy plasma timepoints; longitudinal ctDNA trends captured using a tumor-agnostic WBC DNA informed approach (**A**) are shown alongside dynamic changes in confirmed tumor tissue-derived ctDNA variants (**B**) for a representative patient with mR. Kinetics of tumor-derived mutations identified across serial plasma samples were evaluated by considering the maxMAF of tumor-derived alterations detected at each timepoint. For the assessment of plasma aneuploidy changes (**C**), we examined chromosomal arm-level copy number profiles and utilized the most aberrant chromosomal arm representations in each sample to construct a PA score. Genome-wide arm-level somatic copy number changes for each sampled plasma timepoint are shown alongside plasma aneuploidy (PA) scores in panels C and F. Plasma aneuploidy profiles for the patient shown in A-B mirrored longitudinal changes in maxMAF (**C**). In contrast, the tumor-agnostic WBC-informed approach revealed a pattern of ctDNA persistence for a representative patient with mPD (**D**), that was confirmed by tumor-informed analyses (**E**). Similar to the first patient, PA dynamics mirrored MaxMAF kinetics (**F**), showing persistent cancer-level aneuploidy at all timepoints evaluated. mo, months; OS, overall survival; PFS, progression-free survival; WBC, white blood cell.

### Differential performance of tumor-agnostic WBC DNA-informed vs. tumor-informed and plasma-only approaches

We next assessed the performance of the tumor-agnostic WBC DNA-informed approach in accurately detecting baseline and on-therapy ctDNA using a tumor-informed method as the gold standard. In tandem, we assessed the feasibility of the WBC-informed approach compared to a plasma-only approach, where all cfDNA alterations were considered without an adjudication of cellular origin. These analyses showed that at baseline, a plasma-only cfDNA approach results in a falsely high detectable rate of 96.2% compared to 83.8% when a tumor-agnostic WBC DNA-informed method is employed (**Fig. 5A**). The higher detectable rate of the plasma-only approach is likely driven by contamination by clonal hematopoiesis-related mutations, which are indistinguishable from true tumor-derived alterations. Thus, any baseline samples harboring one such mutation would be falsely labelled as detectable, inflating the overall baseline detection rate in the plasma-only approach.

**Fig 5.**
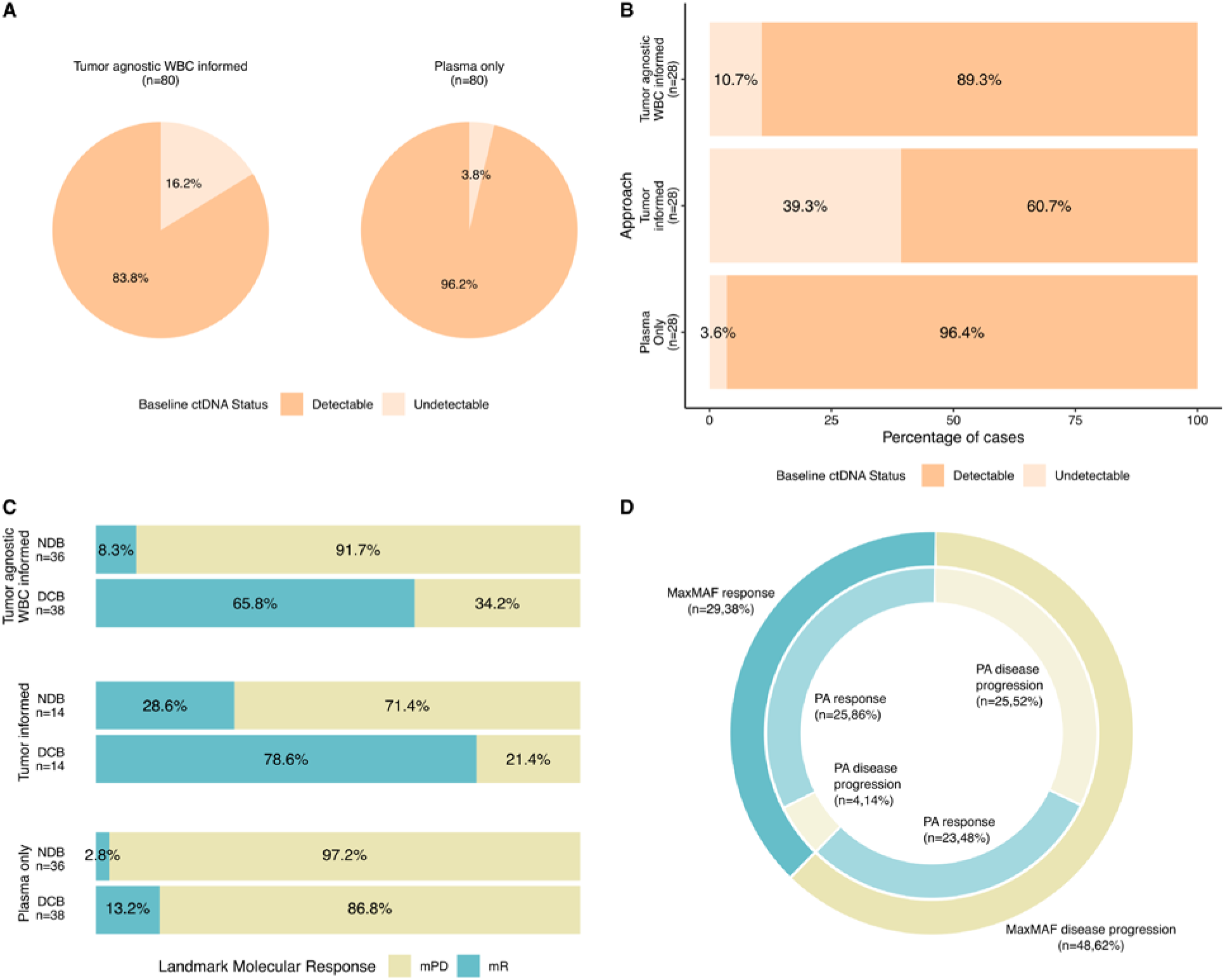
Tumor-agnostic WBC DNA-informed approaches maximise the number of detectable cases at baseline and accurate ctDNA landmark molecular response calls. (**A**) The proportion of patients identified as having detectable ctDNA between baseline and 3-week sampling using our tumor-agnostic WBC DNA-informed variant filtering method was compared to a plasma-only (tumor- and WBC-uninformed) approach. Patients who were not sampled during this interval and those who received durvalumab as maintenance therapy were excluded from analyses. (**B**) Evaluation of the performance of the tumor-agnostic approach, compared to tumor-informed or plasma-only approaches within the subgroup of patients with matched tumor tissue whole exome sequencing. (**C**) Comparative assessment of the accuracy of ctDNA landmark molecular responses using the tumor-agnostic WBC DNA-informed method versus tumor-informed and plasma-only approaches in prediction of durable clinical benefit (PFS ≥6 months). These findings demonstrate that the tumor-agnostic WBC DNA-informed approach maximizes the number of evaluable cases with detectable ctDNA while maintaining the overall accuracy of molecular response assessments by striking a balance between sensitivity (65.8%) and specificity (91.7%) compared to plasma-only (sensitivity 13.2%, specificity 97.2%) or tumor-informed approaches (sensitivity 78.6%, specificity 71.4%). (**D**) Mutant allele fraction (MaxMAF) and longitudinal plasma aneuploidy dynamics were overall concordant in the mR category (p < 0.01, Fisher’s exact test). A moderate degree of concordance was noted between MaxMAF and PA response in the mPD subset. mR, molecular response; mPD, molecular disease progression; PA, plasma aneuploidy; PFS ≥6 months is marked as durable clinical benefit-DCB; PFS <6 months is marked as non-durable clinical benefit-NDB; WBC, white blood cell.

For a subset of patients with available matched tumor tissue whole-exome sequencing (n=28), we conducted a comparison to assess the accuracy of the tumor-agnostic WBC DNA-informed method vs a tumor-informed or plasma-only approach (**Fig. 5B**). As expected, a tumor-informed approach while specific, appeared to be the most stringent and resulted in a higher undetectable rate of 39.3% compared to the WBC-informed approach (10.7%) that in principle is permissive of capturing tumor heterogeneity and evolution (**Fig. 5B**). These findings are line with the notion that in the advanced/metastatic disease setting, the application of a tumor-informed approach is often fraught with challenges arising from the clinical feasibility of tumor sample retrieval, compounded by the limited ability of archival samples to represent the full compendium of somatic alterations due to spatial and temporal tumor heterogeneity. Next, we evaluated the impact of the approach employed in determining landmark molecular response and found that 65.8% and 78.6% of patients with landmark PFS ≥6 months were accurately classified in the mR category by the tumor-agnostic WBC-informed and tumor-informed approaches respectively. With respect to patients with PFS <6months, 91.7% and 71.4% were classified in the molecular progression group respectively (**Fig. 5C**). In contrast, the plasma-only approach showed the poorest performance, correctly classifying only 13.2% of patients with PFS ≥6 months as molecular responders (**Fig. 5C**). Similarly, in subset analyses of the 28 patients analyzed by all three approaches, we observed high concordance (100%) between molecular disease progression and non-durable clinical benefit for both tumor-agnostic WBC-informed and plasma-only approaches compared to the tumor-informed approach (71.4%; fig. S8). More importantly, the plasma-only approach had a strikingly low concordance with durable clinical benefit (14.3%). In comparison, the tumor-agnostic WBC-informed approach had a concordance (71.4%) closer to that of the tumor-informed approach (78.6%; fig. S8). These findings suggest that a tumor-agnostic WBC-informed approach may be more specific in predicting clinical benefit with immunotherapy, while plasma-only approaches are confounded by CH, which renders assessment of molecular response challenging. Importantly, the tumor-agnostic WBC-informed approach increased the number of evaluable cases while maintaining the overall accuracy of molecular response assessments, offering a balance between clinical sensitivity and specificity in this context. In line with the baseline MaxMAF and PA concordance, among the 29 patients with landmark mR, 25 (86%) also demonstrated PA mR. Notably, among the 48 patients in the mPD category, only 25 (52%) were similarly classified by PA, suggesting lower analytical sensitivity and clinical specificity of the PA-only approach in the context of a low circulating tumor fraction (p <0.01, Fisher’s exact test; **Fig. 5D**).

### Clinical sensitivity of ctDNA molecular response in capturing immunotherapy clinical benefit

Despite an increasing body of evidence, a unified definition of molecular response supported by comparative performance of two-timepoint vs landmark assessment is currently lacking. To address this critically important question, we evaluated the clinical sensitivity of landmark ctDNA molecular response (single timepoint assessment, 3-9 weeks on therapy) compared to more traditional molecular response labels that rely on the relative reduction in ctDNA levels between two timepoints (Methods). For two-timepoint molecular response assessments, we considered a tiered approach starting with ctDNA reduction by 50%, followed by increasing incremental reductions by 75%, 90%, 95% and complete clearance (**Fig. 6**). We found that different molecular response assessments yielded variable clinical sensitivity (denoted here as the concordance between ctDNA mR and DCB) and clinical specificity (denoted here as the concordance between ctDNA mPD and NDB) for immunotherapy clinical benefit together with variable fractions of non-evaluable patients when 2 timepoints were required. Single time point determination of landmark molecular response (3-9 weeks) enabled evaluation of the largest fraction of patients (n=74, 100%), while the consideration of maxMAF dynamics (clearance or reduction) resulted in exclusion of a significant fraction (20.3%) of patients with no baseline sample from assessment. Importantly, another 14.9% of patients had undetectable ctDNA across all time points examined, precluding determination of molecular response by consideration of clearance or relative reduction. Notably, in addition to enabling assessment of the largest number of patients, landmark molecular response showed the strongest association with clinical benefit (Fisher’s exact p=2.3e-7), compared to ctDNA clearance (Fisher’s exact p=4.8e-5) or ctDNA reduction of greater than 95% from baseline (Fisher’s exact p=1.6e-4).

**Fig. 6.**
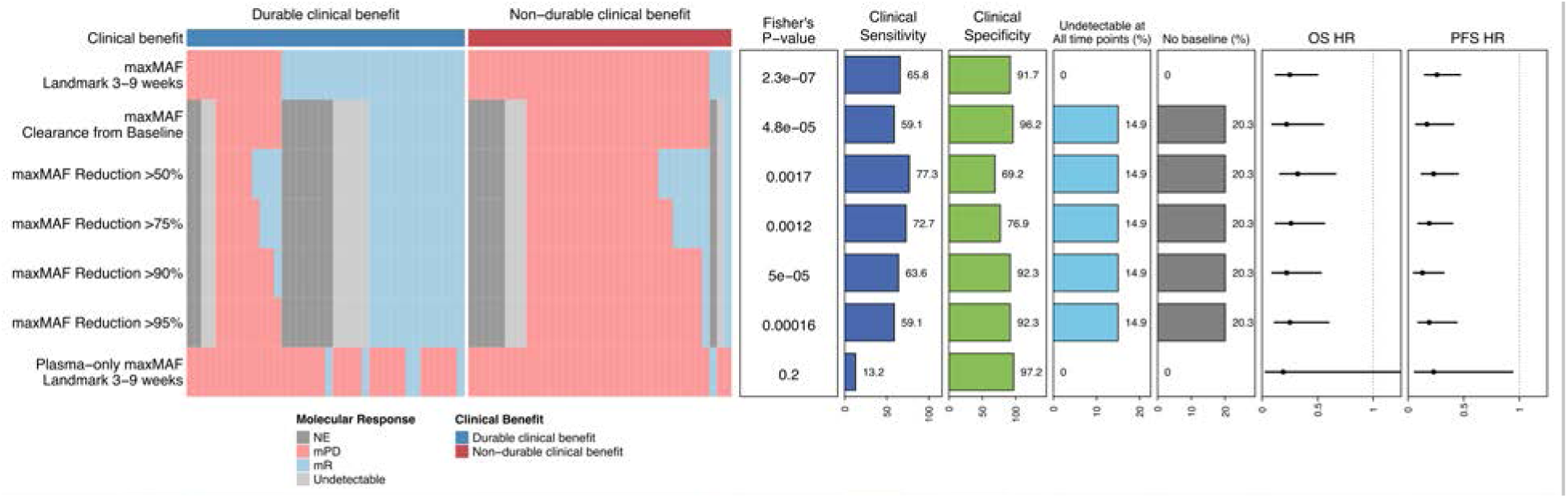
Clinical sensitivity of landmark molecular response in comparison to two-timepoint ctDNA response approaches. The utility of landmark molecular response molecular response in prediction of durable clinical benefit (PFS ≥6 months) is compared to clearance or reduction in ctDNA from baseline. For two-timepoint molecular response definitions, a prediction of ctDNA molecular response could not be made for 35.2% of patients due to unavailability of baseline plasma samples (20.3%) or lack of ctDNA detection at any time point (14.9%), which hinders assessment of clearance or reduction compared to a baseline sample. While maximizing the number of evaluable patients, single timepoint landmark molecular response determined at 3-9 weeks achieved high clinical specificity (91.7%) and sensitivity (65.8%) in predicting clinical benefit while showing similar performance to two-timepoint approaches in prediction of progression-free (landmark, HR=0.26 [95% CI 0.15-0.47], logrank p=1.6e-06; clearance from baseline, HR=0.17 [95% CI 0.07-0.41], logrank p=1.1e-05; reduction>95% from baseline, HR=0.19 [95% CI 0.09-0.44], logrank p=1.7e-05) and overall survival (landmark, HR=0.25 [95% CI 0.12-0.5], logrank p=2.5e-05; clearance from baseline, HR=0.22 [95% CI 0.09-0.55], logrank p=4.7e-4; reduction>95% from baseline, HR=0.25 [95% CI 0.11-0.6], logrank p=9.2e-4). In contrast to tumor agnostic WBC informed assessment using any single (landmark, Fisher’s exact p=2.3e-7) or two timepoint definitions (clearance from baseline, Fisher’s exact p=4.8e-6; >95% reduction from baseline, Fisher’s exact p=1.6e-4) of molecular response, no significant association with clinical benefit (clinical benefit, Fisher’s exact p=0.20) or overall survival (HR=0.19 [95% CI 0.03-1.37], logrank p=0.064) and a weak association with progression-free survival (HR= 0.23 [95% CI 0.06-0.94], logrank p= 0.026) is noted for the landmark molecular response based on the plasma only approach. Clinical sensitivity is defined as the fraction of patients with durable clinical benefit that have concordant molecular response assignment, and clinical specificity indicates the fraction of patients with non-durable clinical benefit that have a concordant molecular progressive disease assignment. Fisher’s exact test is used to evaluate the association between each molecular response measure and clinical benefit. NE, not evaluable; mPD, molecular progressive disease; mR, molecular response; OS, overall survival; PFS, progression free survival; HR, hazard ratio.

Tying ctDNA molecular response into clinical outcomes, landmark molecular response was significantly associated with longer overall (OS) and progression-free survival (PFS) across different approaches with a similar range of associated hazard ratios (OS HR range 0.22-32, PFS HR range 0.13-0.26). To evaluate the contribution of matched WBC analysis in the determination of molecular response, we also implemented a plasma-only approach to define molecular response at the 3-9 landmark interval. The plasma-only landmark molecular response assessment achieved the lowest clinical sensitivity among all approaches (13.2%), highlighting the challenges posed by biological noise due to clonal hematopoiesis. To better elucidate the accuracy of the tumor-agnostic WBC-informed approach in assessment of molecular response, we compared the performance of this approach to the tumor-informed approach in the subset of patients with matched tumor exome data (n=28, Fig. S9). In this subset, among patients with durable clinical benefit, landmark assessment of molecular response resulted in similar predictions of response using tumor-informed (clinical sensitivity of 78.6%) and tumor-agnostic WBC-informed (clinical sensitivity of 71.4%) approaches. For patients with non-durable clinical benefit, the tumor agnostic WBC approach accurately predicted lack of response in all cases (clinical specificity of 100%), while the tumor-informed approach resulted in discordant predictions for 4 out of 14 patients (clinical specificity of 71.4%). No significant association with clinical benefit was noted for the plasma-only approach (Fisher’s exact p=0.48). Taken together, these findings suggest that landmark ctDNA assessment 3-9 weeks on immunotherapy has reasonable sensitivity and specificity and captures a larger number of patients, as such representing a credible approach as an early endpoint of immunotherapy response.

### Evaluation of the heterogeneity and clinical value of ctDNA detection dynamics within the landmark window

To further rationalize our definition of landmark molecular response, we evaluated the heterogeneity of ctDNA detection within the landmark interval and assessed the impact of landmark interval dynamics on progression-free survival (**Fig.7**). Of the 77 evaluable patients within the landmark interval, 43 (55.8%) had two or more samples collected between 3 and 9 weeks after treatment initiation. Of this subset, 31 patients (72.1%) had the same ctDNA detection status across all analyzed plasma samples, with 23 consistently detected and 8 consistently undetected. The remaining 12 patients (27.9%) switched from detected to undetected (n=7) or vice versa (n=5; **Fig.7A**). Patients with consistent ctDNA detection across the landmark interval had similar progression-free survival to those with a single available plasma sample with detectable ctDNA (n=23 vs 25, median PFS=3.9 vs 3.1 months, logrank p=0.81; **Fig.7B**). Similarly, patients with consistently undetected ctDNA in the landmark interval (n=8) did not have statistically significantly longer PFS than those with a single available plasma sample with undetectable ctDNA (n=9, logrank p=0.23). Notably, patients with variable ctDNA detection status within the landmark interval, had a significantly longer PFS compared to those with consistent or singular detected ctDNA (n=12 vs 48, median PFS=26.6 vs 3.4, logrank p=0.0005), and similar to those with consistent or singular undetected ctDNA (n=12 vs 17, median FPS=26.6 vs 24.3, logrank p=0.82; **Fig.7B**). Therefore, we reasoned that, given the observed differences in progression-free survival, patients with absent ctDNA at at least one plasma timepoint within the landmark interval constitute a distinct clinical entity with outcomes comparable to those of patients in whom ctDNA remains detectable across all available plasma samples.

**Fig. 7.**
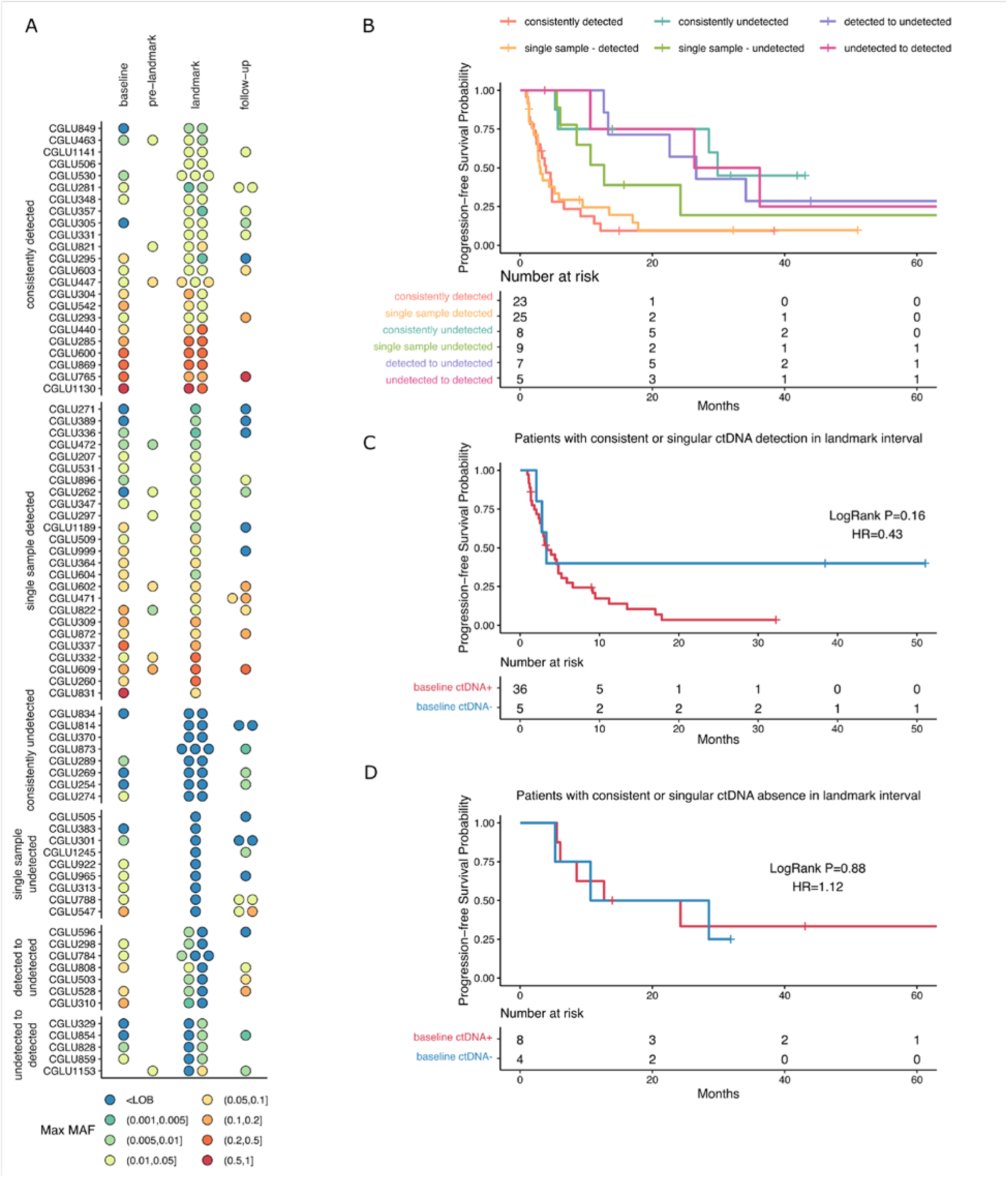
Association of dynamics of ctDNA detection with clinical outcomes. **A.** Available plasma timepoints from each patient are classified as baseline (prior to treatment initiation), pre-landmark (within 3 weeks following treatment initiation), landmark (3-9 weeks from treatment initiation), and follow-up interval, and the maximum mutant allele fraction (Max MAF) at each time is displayed. For patients with more than one plasma collection per interval, the Max MAF values are ordered sequentially within the interval column from left (earlier) to right (later). Dynamic changes In ctDNA detection within the landmark interval is used to define six distinct categories of patients (top to bottom): [1] patients with consistent ctDNA detection in more than one timepoint. [2] patients with one available plasma sample within landmark interval that is positive for ctDNA. [3] patients with consistent absence of ctDNA in more than one timepoint. [4] patients with one available plasma sample within landmark interval that is negative for ctDNA. [5] patients where ctDNA is detected early in the landmark interval but is absent in later samples of the same interval. [6] patients with absent ctDNA early in the landmark interval followed by ctDNA detection. **B.** ctDNA dynamics predict distinct patterns of progression-free survival. Patients with consistent detection of ctDNA in the landmark interval (with more than one plasma sample) had a similar progression-free survival to those with a single available plasma sample with detectable ctDNA (n=23 vs 25, median PFS=3.9 vs 3.1 months, logrank p=0.81). Median progression-free survival was longer in patient with consistent absence of ctDNA in landmark interval (n=8, 29.9 months) compared to those with a single available plasma sample with undetectable ctDNA (n=9, 12.7 months), but the difference was not statistically significant (logrank p=0.23). Patients with variable detection status had a significantly longer PFS compared to those with consistent or singular detection of ctDNA within the landmark interval (n=12 vs 48, median PFS=26.6 vs 3.4, logrank p=0.0005), but similar to those with consistent or singular absent ctDNA (n=12 vs 17, median FPS=26.6 vs 24.3, logrank p=0.82). **C.** Among patients with consistent or singular detection of ctDNA in the landmark interval, ctDNA detection at baseline did not correlate with a significant difference in progression-free survival (detectable vs undetectable, n=36 vs 5, median PFS=3.4 vs 3.3 months, logrank p=0.16). **D.** Similarly, baseline detection status did not result in different PFS among patients with consistent or singular undetectable ctDNA at landmark (detectable vs undetectable, n=8 vs 4, median PFS=18.5 vs 19.6, logrank p=0.88).

Next, we evaluated the impact of baseline ctDNA detection while accounting for landmark interval ctDNA status. Among patients with consistent or singular detectable ctDNA at landmark, those with (n=36) or without (n=5) detectable ctDNA at baseline had similar PFS (median PFS=3.4 vs 3.3 months, logrank p=0.16, **Fig.7C**). Similarly, baseline ctDNA detection status did not result in different PFS among patients with consistent or singular undetectable ctDNA at landmark (detectable vs undetectable, n=8 vs 4, median PFS=18.5 vs 19.6, logrank p=0.88, **Fig.7D**). The lack of additional survival stratification by baseline detectability after accounting for landmark status further supports the notion that landmark ctDNA molecular response sufficiently prognosticates survival outcomes and circumvents the logistical and financial burden of the alternative two timepoint assessment paradigm.

### Landmark molecular response independently predicts progression-free and overall survival

We found a significant association between landmark ctDNA molecular response and PFS ≥6 months for all patients (p=2.3e-07, Fisher’s exact test; **Fig. 8A**) as well as in patients receiving single-agent immunotherapy (p=2.5e-05, Fisher’s exact test) or chemo-immunotherapy (p=0.02, Fisher’s exact test; fig. S10). Specifically, landmark ctDNA molecular response was highly predictive of PFS ≥6 months across all patients (positive predictive value; PPV=0.89), those receiving single-agent immunotherapy (PPV=0.89) or chemo-immunotherapy (PPV=0.90). Importantly, associations between molecular responses and survival outcomes revealed that patients with mPD had significantly shorter PFS (median 3.42 vs 26.63 months, logrank p=1.6e-06; **Fig. 8B**) and OS (median 11.38 vs 46.95 months, logrank p=2.5e-05; fig. S10) compared to patients who attained mR. Similar findings we encountered in subset analyses of patients receiving IO monotherapy with respect to PFS (median 3.32 vs 29.9 months, logrank p=3e-05; **Fig. 8C-D**) and OS (median 11.34 months vs not reached-NR, logrank p<0.001; fig. S10). In the subset of patients treated with combination chemo-IO, there was a significant difference in progression-free survival between the two response groups (mPD vs mR, median PFS 4.4 vs 12.7 months, logrank P=0.05; fig. S10D), while a numeric difference in overall survival was noted (mPD vs mR, median OS 11.4 vs 33.0 months, logrank P=0.096; fig. S10C); the reduced statistical significance may be attributed to the comparatively smaller number of patients in the chemo-IO subset. Evaluation of landmark molecular responses across histological subtypes revealed consistent associations with clinical benefit and survival independent of adenocarcinoma or squamous cell carcinoma histology (fig. S11).

**Fig. 8.**
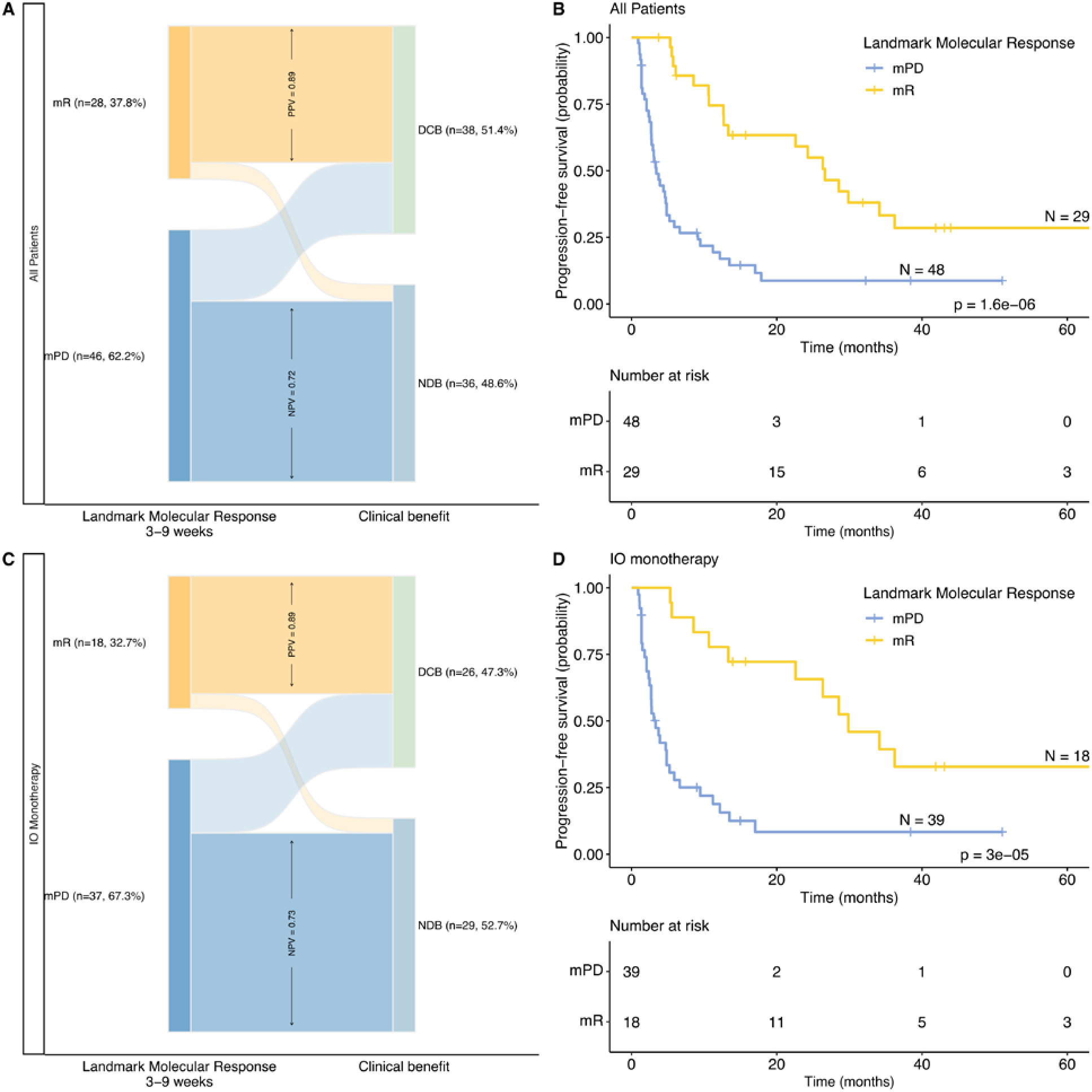
Molecular response is associated with durable clinical outcomes with immunotherapy. (**A**) ctDNA molecular responses were significantly associated with landmark PFS ≥6 months (marked as durable clinical benefit-DCB in all evaluable study patients (P < 0.001, Fisher’s exact test). (**B**) Patients with early on-therapy molecular response (mR) had a significantly longer progression-free survival (PFS) compared to patients with molecular progressive disease (mPD). (**C**) A similar correlation between ctDNA landmark molecular response and clinical benefit was noted in the subset of patients treated with single-agent immunotherapy (P < 0.001). Molecular disease progression had moderate predictive power for short PFS (PFS<6 months; negative predictive value-NPV; all patients NPV=0.72, IO monotherapy NPV=0.73, chemo-IO NPV=0.67). (**D**) A similar association between early on-therapy molecular response and longer progression-free survival was observed in the subset of patients receiving single-agent immunotherapy. mR, molecular response; mPD, molecular disease progression; PFS <6 months is marked as non-durable clinical benefit-NDB.

The association between landmark molecular response and PFS (HR=0.26, 95% CI=0.14-0.48, p <0.001) and OS (HR=0.23, 95% CI=0.11-0.49, p <0.001) remained statistically significant after accounting for clinical covariates and line of IO therapy in a multivariate Cox proportional hazard regression model (**Fig. 9A-D**, tables S13-14). Importantly, landmark ctDNA molecular response independently predicted OS and PFS when the baseline circulating tumor fraction was also considered, highlighting the value of molecular response across the spectrum of tumor fraction and in cases with a tumor fraction <1% (overall survival, HR=0.19, 95% CI=0.07-0.47, p<0.001, **Fig. 9B**; progression-free survival, HR=0.26, 95% CI=0.13-0.53, p<0.001, **Fig. 9D**). We considered additional assessments of baseline ctDNA levels, including continuous and dichotomized by the median values; these analyses revealed that there was no significant association between continuous baseline maxMAF values and progression-free (HR=0.51 [95% CI 0.04-6.90], p=0.61) or overall survival (HR=0.69 [95% CI 0.04-12.58], p=0.80), while landmark molecular response remained a significant predictor (HR=0.24 [95% CI 0.12-0.50], p<0.001; HR=0.19 [95% CI 0.07-0.51], p=0.001 for PFS and OS respectively). Multivariate Cox models including a dichotomized baseline maxMAF covariate (using the cohort median) showed a significant association between PFS and OS and both landmark molecular response and baseline maxMAF. Still, the association was stronger for landmark molecular response (PFS, molecular response HR=0.28 [95% CI 0.14-0.56], p<0.001, baseline maxMAF HR=0.51 [95% CI 0.26-0.97], p=0.042; OS, molecular response HR=0.21 [95% CI 0.08-0.52], p=0.001, baseline maxMAF HR=0.37 [95% CI 0.17-0.83], p=0.015; fig. S12). Taken together, our findings elucidate the technical and biological ramifications of landmark ctDNA molecular response together with the clinical value of ctDNA responses as an early predictor of immunotherapy response for patients with metastatic NSCLC.

**Fig. 9.**
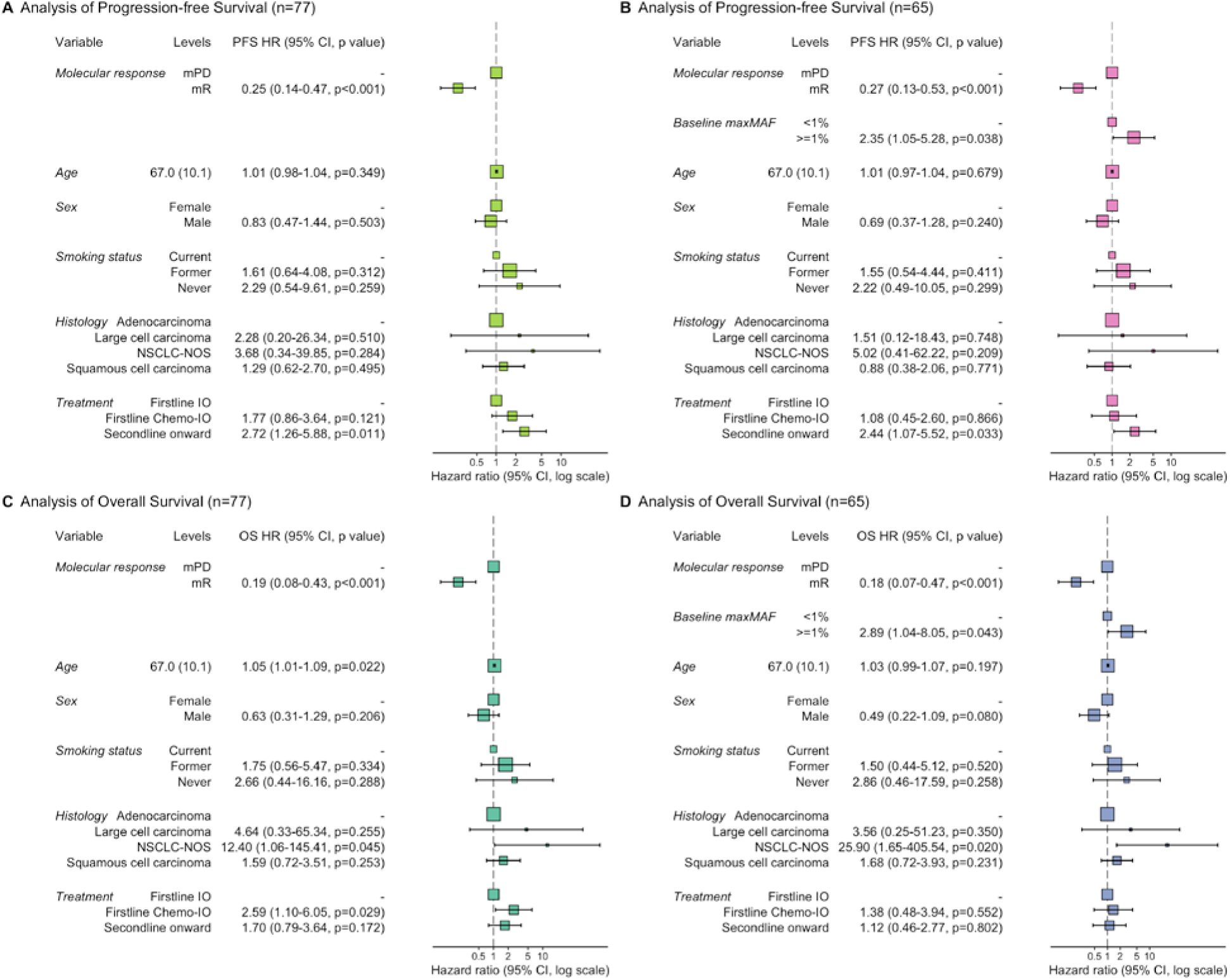
Independent prediction of progression-free and overall survival for landmark molecular response. Multivariate Cox proportional hazards regression analyses showed a significant independent association between landmark ctDNA molecular response and progression-free (**A,** HR=0.25 [95% CI 0.14-0.47], p<0.001; B HR=0.27 [95% CI 0.13-0.53], p<0.001) and overall survival (C, HR=0.19 [95% CI 0.08-0.43], p<0.001; D, HR=0.18 [95% CI 0.07-0.47], p<0.001) after accounting for clinical covariates including sex, smoking status, histology, and treatment regimen. This association remained significant after inclusion of baseline ctDNA fraction, dichotomized at 1% maxMAF value, as an independent predictor (**B, D**). Patients receiving first line immunotherapy or chemo-immunotherapy had longer progression-free survival compared to patients on second (or higher order) lines of therapy (second line onward; A, HR=2.72 [95% CI 1.26-5.88], p=0.011; B, HR=2.44 [95% CI 1.07-5.52], p=0.033). In the multivariate models, no significant association was observed between overall survival and the clinical covariates including sex, smoking status. The weak association between overall survival and NSCLC-NOS histology was likely spurious due to the small sample count (n=1), while other histologies did not show a significant association. For each clinical or genomic covariate, hazard ratios compared to the reference group, along with 95% confidence intervals and the associated p-value (Wald test) are listed and displayed in the forest plot.

## Discussion

There is an urgent need to establish a standardized framework to assess the utility of liquid biopsies and ctDNA molecular responses in monitoring response to ICI therapy among patients with advanced/metastatic NSCLC. While the clinical relevance of ctDNA molecular response in predicting therapeutic outcomes has been previously reported (10, 12, 16, 21, 22) including in a clinical trial setting (13), further studies are needed to solidify the definition, timing and clinical sensitivity of ctDNA molecular response and bridge these advancements with routine clinical care.

We demonstrated that landmark ctDNA molecular response, assessed 3-9 weeks from ICI initiation, in a tumor-agnostic WBC-informed manner can identify patients at risk of disease progression, in whom early intervention and treatment escalation and/or prompt therapeutic switching may manifest a meaningful improvement in outcome. Currently, considerable differences exist in the definition of ctDNA molecular response across studies, which are largely shaped by assay-specific variations in sensitivity and negative predictive value (8). Building on the results from previous proof-of-concept studies (10, 22–24) and the first stage of the BR.36 clinical trial (NCT04093167) (13), that prospectively assessed the timing, definition and concordance of ctDNA response with radiographic response, we utilized single time point landmark molecular response and showed that landmark molecular response serves as a predictor for both clinical benefit and long-term survival outcomes. We posit that landmark molecular response maximizes clinical sensitivity of the approach together with the fraction of evaluable patients. Single timepoint assessment, analogous to the post-operative minimal residual disease setting, represents a clinically feasible approach and can be used as a molecular readout of immunotherapy response in advanced stage or metastatic settings. Importantly, this approach circumvents the added financial and logistical burden associated with collection and analysis of multiple plasma timepoints, and enables evaluation of patients where baseline plasma samples are unavailable (which accounted for 20% of patients in the current cohort). This concept is now tested in a ctDNA-adaptive clinical trial setting, in the second stage of the BR36 trial, where patients with landmark ctDNA mPD 6-9 weeks after pembrolizumab are randomized to either pembrolizumab continuation or therapy optimization with pembrolizumab and chemotherapy (NCT04093167) (13).

While various metrics, such as mean (11) or maximal MAF (12, 16, 21, 25) of tumor-derived alterations, have been proposed to quantify dynamic circulating tumor fraction changes during immunotherapy, a consensus on circulating tumor fraction definition remains elusive to date. Recent studies and pooled analyses have echoed our findings that changes in maximal mutant allele frequency of tumor-derived variants are most indicative of therapeutic response (11). While the added value of integrative approaches leveraging both sequence and structural alterations for circulating cell-free tumor load tracking is less studied (16, 26, 27), multi-feature modelling of tumor fraction may increase the sensitivity of ctDNA detection. Nevertheless, assessment of plasma aneuploidy, derived from on-target and off-target sequence data from hybrid capture cfDNA next-generation sequencing, while highly concordant with mutation-based tumor fraction estimates, has a lower analytical sensitivity (28). More specifically, the sensitivity of plasma aneuploidy assessments may be limited by susceptibility to technical limitations which highlight the challenges with accurate tumor fraction estimation at the lower end of the spectrum (26, 27). Consistent with this notion, we found a high concordance between MaxMAF and PA molecular response, but a less tight association between mPD and PA progression.

Furthermore, navigating between a tumor-informed liquid biopsy approach, that conceptually maximizes sensitivity but is limited by feasibility, and a tumor-agnostic strategy represents a challenging task and a critical clinical question. Furthermore, in the metastatic setting, access to tumor tissue is frequently challenging, and available tumor samples do not capture the full set of somatic alterations present across all metastatic sites due to tumor evolution and intratumor heterogeneity. These challenges reflect the practical limitations of applying a tumor-informed liquid biopsy approach in the metastatic setting, which currently preclude its clinical scalability. In this study, when compared to a tumor-informed strategy, the tumor-agnostic WBC DNA-informed approach maintained the accuracy of molecular response calls, supporting its validity. WBC DNA-informed strategies can enhance the specificity of cell-free tumor load monitoring compared to plasma-only methods, while still capturing tumor diversity and filtering out biological noise from mutations associated with CH (29, 30). In line with this notion, we found that the tumor-agnostic WBC DNA-informed approach strikes a balance between sensitivity and specificity, accounts for non-tumor cfDNA mutations, as well as tumor heterogeneity and evolution.

A critical consideration in assessing the value of ctDNA molecular response is the evaluation of cases where ctDNA levels are low and there is uncertainty about the accuracy of ctDNA in capturing clinical outcomes. To this end, we included baseline ctDNA levels in multivariable modeling to adjust for cfDNA tumor fraction, ensuring that the predictive value of landmark molecular response for PFS and OS remains independent of whether baseline ctDNA levels are high or low. Indeed, continuous or dichotomized maxMAF values did not affect the independent predictive role of landmark ctDNA response. These findings suggest that the landmark molecular response is independent of the baseline ctDNA level, an important conclusion given the challenges of precise molecular response assessment when ctDNA levels are low, particularly near the limit of detection of the liquid biopsy assay.

In addition to monitoring early on-therapy molecular responses, we demonstrated the value of longitudinal molecular genotyping of NSCLC during ICI. While cfDNA comprehensive molecular profiling has been established for detection of actionable mutations, matching with targeted therapies and tracking emergence of resistance mutations (31), longitudinal plasma genomic profiling is not routinely performed to capture the evolving genomic landscape in the context of immunotherapy. Our findings suggest that the evolving ctDNA landscape has the potential to capture genomic determinants of ICI response which may be used to assist treatment decisions. With a faster turnaround time and elimination of the need for invasive tissue sampling, ctDNA-based molecular genotyping may provide a valuable alternative to tissue testing, particularly in patients who are not fit to undergo invasive tissue sampling or when available tissue biopsies are inadequate for molecular analysis due to quantity or tumor purity. In addition, we demonstrated the utility of ctDNA assessments to dissect the differential genomic profiles of NSCLC histological subtypes, which may provide a valuable tool to aid clinical decision-making.

Limitations of our study include the heterogeneity of blood sampling and treatment regimens. Moreover, RECIST assessments were not available for this real-world cohort. Despite the significant association between landmark molecular response and survival outcomes, a fraction of patients who attained PFS ≥6 months in our study did not attain a molecular response. We believe that this was attributed due to sampling variability that allowed sampling as early as 3 weeks after ICI initiation.

Taken together, we show that landmark ctDNA molecular response has the potential to be developed as an early endpoint of immunotherapy response. Assessing landmark ctDNA molecular response 3-9 weeks after ICI initiation, provides a key window of opportunity for early interception of primary therapy resistance with the timely application of treatment intensification and/or switching strategies.

## Data Availability

All data produced in the present work are contained in the manuscript

## Data availability

Targeted next-generation sequence data are deposited and can be retrieved from the European Genome-Phenome Archive (EGA study accession EGAS_pending, EGA dataset accession EGAD_pending). All computational pipelines used in this work are explicitly described and referenced in the methods section. No custom code was used for this work.

## Author contributions

Conceptualization: LS, NN, PF, VA, Methodology: LS, NN, AB, CC, VA, Investigation: JM, KM, JB, PF, VA, Visualization: JW, Funding acquisition: JB, PF, VV, VA, Project administration: GP, SN, CB, IB, TL, Supervision: VA, Writing – original draft: LS, NN, VA

## Acknowledgements

We would like to thank the clinical research personnel at the Johns Hopkins Upper Aerodigestive Malignancies Program for their contributions with patient enrolment and biospecimen procurement.

