## Supplementary Figures S1-S12 for "Landmark ctDNA molecular response represents an early predictor of immunotherapy outcomes in lung cancer: A clinical utility study"

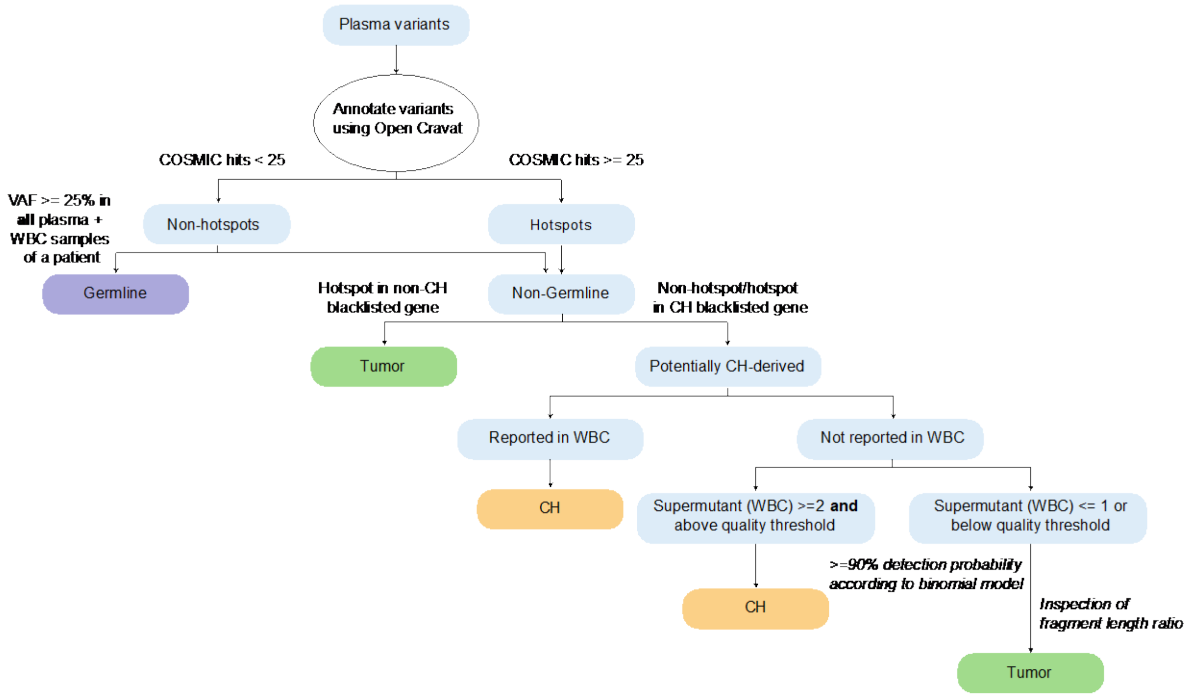


**Figure S1. Overview of branched logic architecture used for classification of variant cellular origin.** Plasma variants were initially cross-referenced against the Catalogue of Somatic Mutations in Cancer (COSMIC) v95 database for hotspot alteration annotation, utilizing OpenCRAVAT. A conservative COSMIC frequency threshold of 25 hits was applied to define a cancer hotspot. Variants not classified as hotspots, yet exhibiting a VAF >25% in all plasma and available WBC samples from a patient, were designated as germline. Plasma variants detected in blacklisted genes commonly altered in CH, with at least 2 supporting supermutants above assay-specific quality thresholds in matched WBC TEC-Seq data were considered CH-derived, provided they were not lung cancer hotspots. Plasma variants not detected in matched WBC sequence data (supermutant count <2 or below assay-specific quality thresholds) were classified as follows: for each assessed genomic position, the binomial distribution probability was calculated using the observed distinct coverage for the position, the number of observed mutant reads, and the corresponding variant allele fraction. Alterations with a detection probability $\geq$90% underwent visual inspection to further exclude likely sequencing and mapping artifacts, and the length ratio of mutant to wild-type fragments was assessed. Alterations passing quality inspection were classified as tumor-derived.

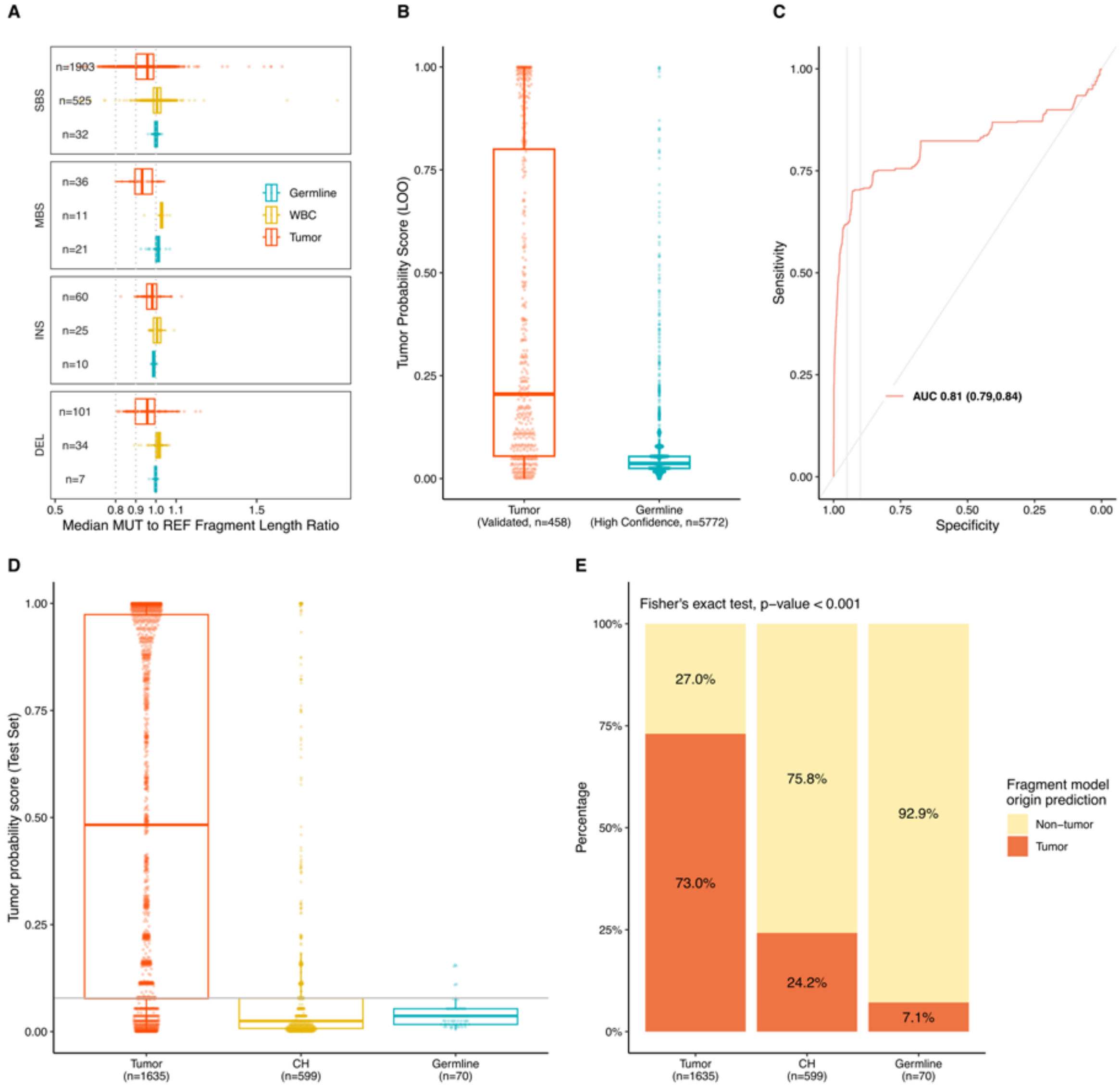


**Figure S2. Assessment of variant cellular origin using median fragment length ratios.** (**A**) For each germline, clonal hematopoiesis (CH), or tumor-derived variant, we calculated the ratio (r) between the median length of fragments harboring the mutant allele compared to the wild-type allele independent of mutation type (i.e., substitution, insertion, deletion, or multi-base substitution). For germline or CH variants, the mutant and wild-type fragments had a similar median length (germline mean r = 1.00, range 0.98-1.03; CH mean r = 1.00, range 0.54-1.90), while for tumor-derived variants, mutant fragments were slightly shorter compared to wild-type fragments (mean r = 0.95, range 0.56-1.63). Given these differences, we reasoned that length differences between fragments with mutant or wild-type alleles could be used to inform variant origin. (**B**) We built a logistic regression model on a training set comprised of bona fide germline variants (n=5772), and tumor-derived mutations (n=458). Germline variants were selected by analysis of sequence alterations in WBC samples and were required to be at a polymorphic site listed in the dbSNP database (build ID 151), pass all quality filters, and have mutant allele fraction of 40-60% in the WBC sample. Tumor-derived mutations were those confirmed to be somatic alterations by analysis of whole exome sequencing data of the tumor sample from the same individual. For each mutation, the model predicts the probability of being tumor-derived given the mutant to wildtype median fragment length ratio. We implemented a nested LOO cross-validation training setup. In each iteration, all plasma variants from a given patient were set aside (outer loop), and the parameters of the logistic regression model were estimated using a 10x5 training setup (10 repeats of 5 fold CV) in the inner loop. The final model resulting from the inner loops training was used to generate the probability scores for the hold-out plasma variants (outer loop), offering an unbiased estimate of the model's performance in previously unseen new variants. (**C**) Using the ground-truth variant origin labels for the hold-out plasma variants (in LOO setup), we evaluated the model performance by ROC analysis (AUC 0.81, 95% CI 0.79 – 0.84). (**D**) For all reported plasma variants outside the training set, the logistic regression model predicted the probability of being tumor-derived. We also calculated the score threshold corresponding to a specificity of 90% coupled with a sensitivity of 70% in the training set (threshold probability score > 0.0783). The threshold probability score is indicated by the dotted line. (**E**) At the selected model parameters, 73% tumor-derived variants were concordantly classified based on fragment length. In addition, the majority of germline (93%) and CH (76%) alterations were concordantly identified as non-tumor origin based on median fragment length ratios, underscoring the utility of fragment length profiling as a complementary tool to evaluate the specificity of variant origin calls in plasma. The analyses were performed in R v.4.3, using packages caret (v.6.0-94) and pROC (v.1.18.4). SBS, single base substitution; MBS multi-base substitution; INS, insertion; DEL, deletion.

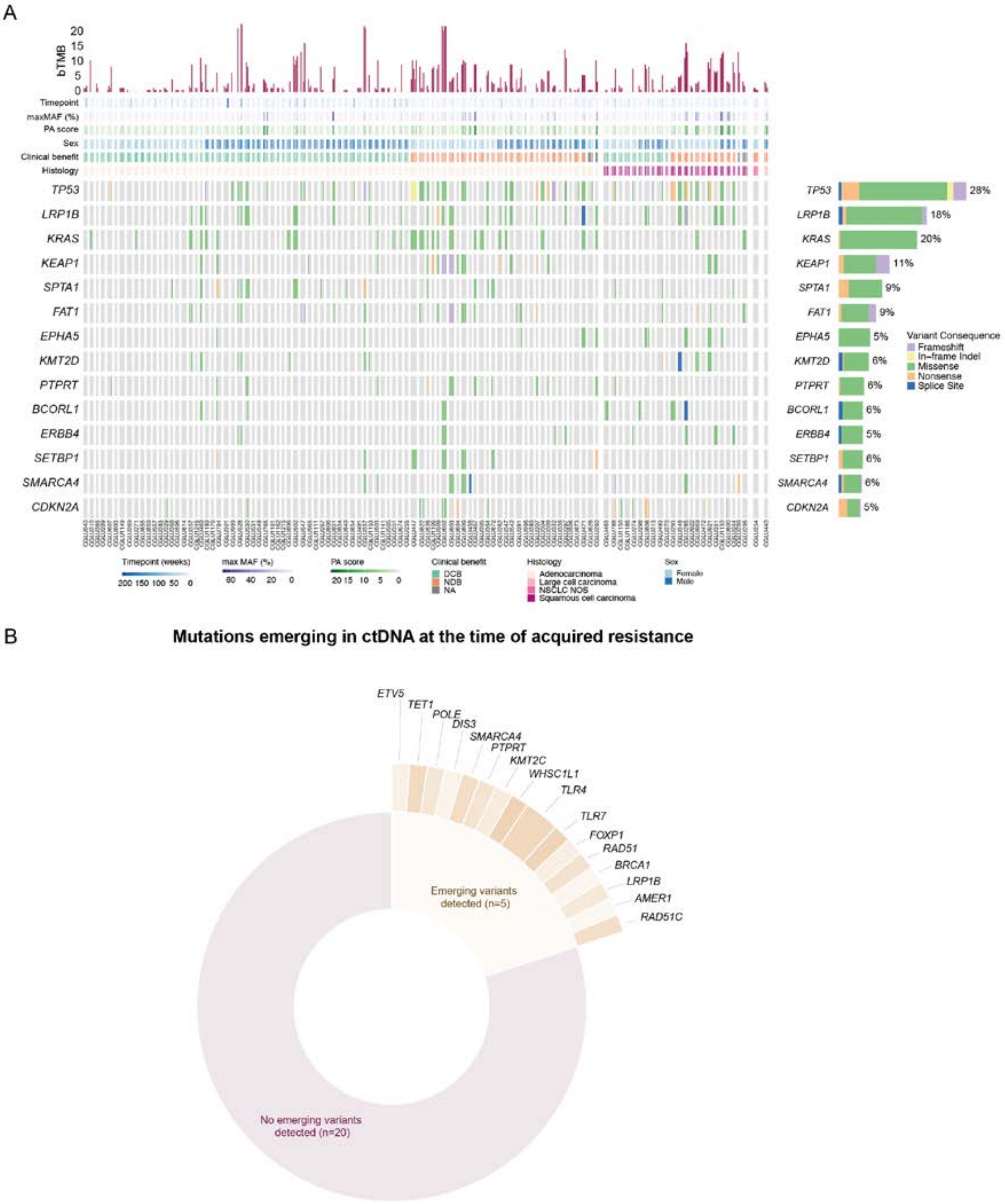


**Figure S3. Landscape of baseline and emerging ctDNA mutations at the time of molecular disease progression.** (**A**) Overview of the landscape of tumor-derived ctDNA mutations detected in plasma. The frequencies of mutations at the sample level for each gene are displayed on the right and blood tumor mutation burden (bTMB) measurements from each sample are depicted on the top bar plot. Patients attaining progression-free survival (PFS) ≥6 months are indicated as durable clinical benefit (DCB) while patients with a PFS <6 months are annotated as attaining non-durable clinical benefit (NDB). (**B**) Out of the 25 patients identified as molecular responders in the study cohort, five developed acquired resistance to immunotherapy and had newly emerging mutations in ctDNA coinciding with disease progression.

Mutations that were newly acquired in ctDNA at the time of molecular progression were verified against available matched tumor tissue WES and/or clinical tumor molecular profiling data abstracted from electronic health records to confirm absence at baseline. Emerging mutation MAFs ranged from 0.60 to 8.51%. Pie-donut plot displaying genes that harbored newly emerging ctDNA variants at disease progression.

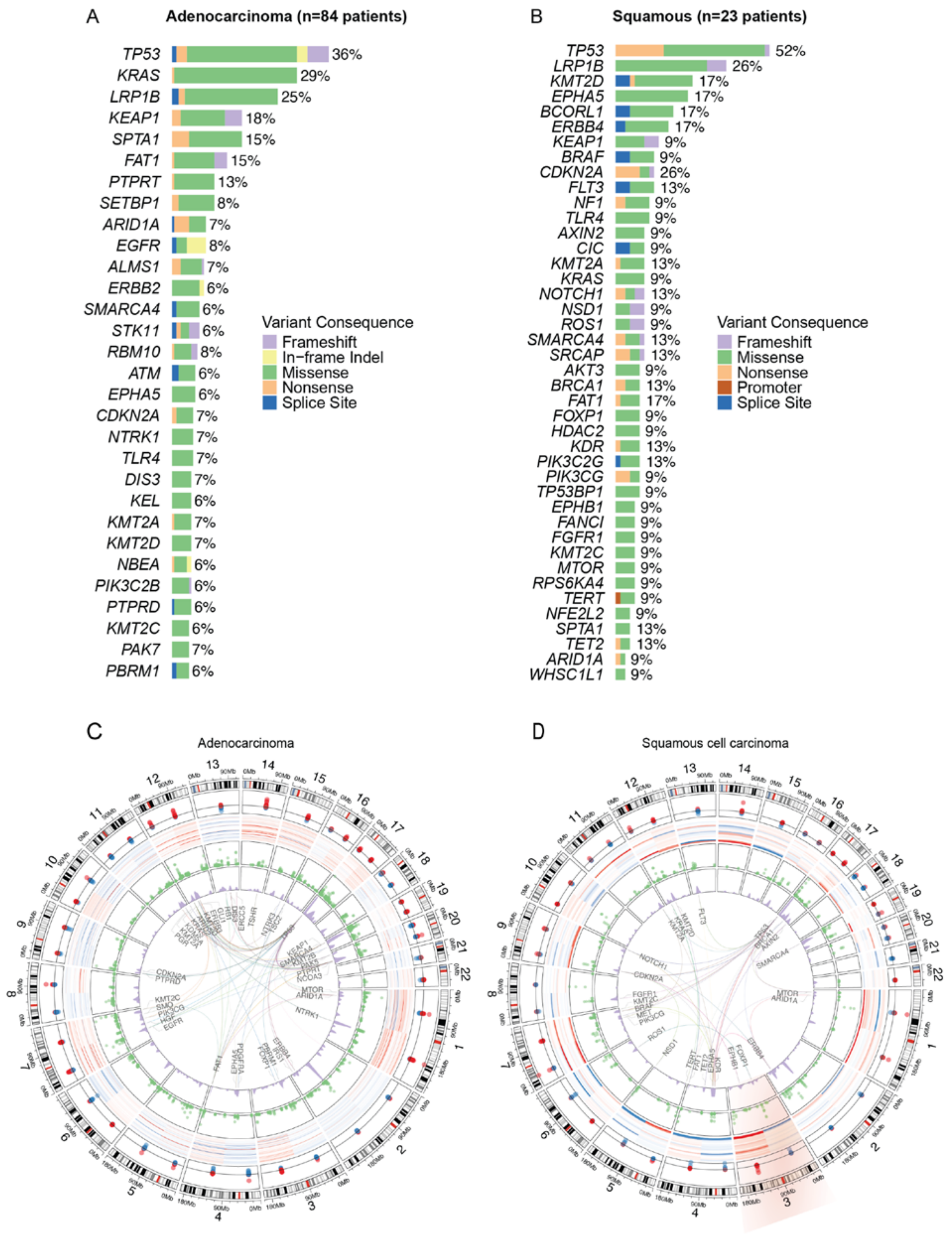


**Figure S4. Recurring sequence and structural genomic alterations by NSCLC histologic subtype.** Genes harboring ctDNA mutations at >5% prevalence amongst patients with (**A**) adenocarcinoma and (**B**)

squamous subtype tumors from the study cohort are shown. The frequency of individual gene alterations within each respective subtype is indicated to the right of each bar. (**C-D**) Circos plots depicting sequence and structural alterations identified in ctDNA according to histology: patients with adenocarcinoma subtype NSCLC tumors are shown in (**C**) and patients with squamous subtype tumors are shown in (**D**). Chromosomal arm-level z scores aggregated from baseline plasma samples are shown on the outer tracks, where gains in copy number are indicated in red and copy number losses are indicated in blue. The variant allele fraction distribution of tumor-derived ctDNA alterations is shown in track three (green) and the density of ctDNA mutations detected across chromosomal regions within each subgroup is shown in the fourth track (purple). Frequent co-occurring mutations (present in >2 distinct patients) in each subtype are shown in the centre.

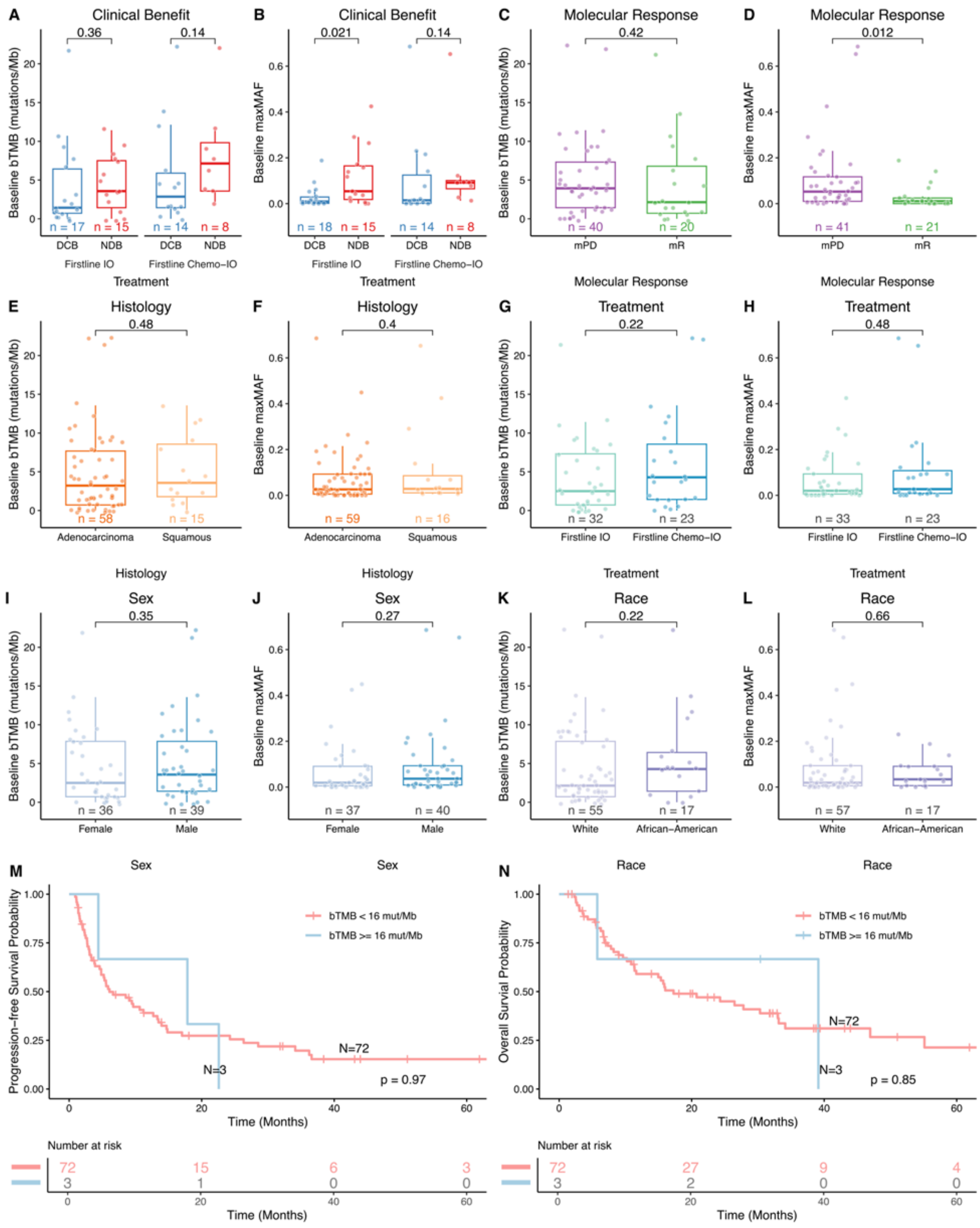


**Figure S5. Association between baseline ctDNA features and clinicopathological characteristics.** Associations between the maximum mutant allele fraction (maxMAF) of tumor-derived alterations and blood tumor mutation burden (bTMB) at baseline timepoints and (**A-B**) progression-free survival (PFS ≥6

months; durable clinical benefit-DCB or PFS <6 months; non-durable clinical benefit-NDB), (**C-D**) molecular response, (**E-F**) histology, (**G-H**) treatment, (**I-J**) sex and (**K-L**) race are shown. Mann-Whitney U test is used to determine the statistical significance of the observed differences between groups and the p-values are indicated. Baseline bTMB measurements above or below 16 mutations/Mb did not show a significant association with (**M**) PFS (log-rank p = 0.97) or (**N**) OS (log-rank p = 0.85).
Abbreviations; mR, molecular response; mPD, molecular progressive disease.

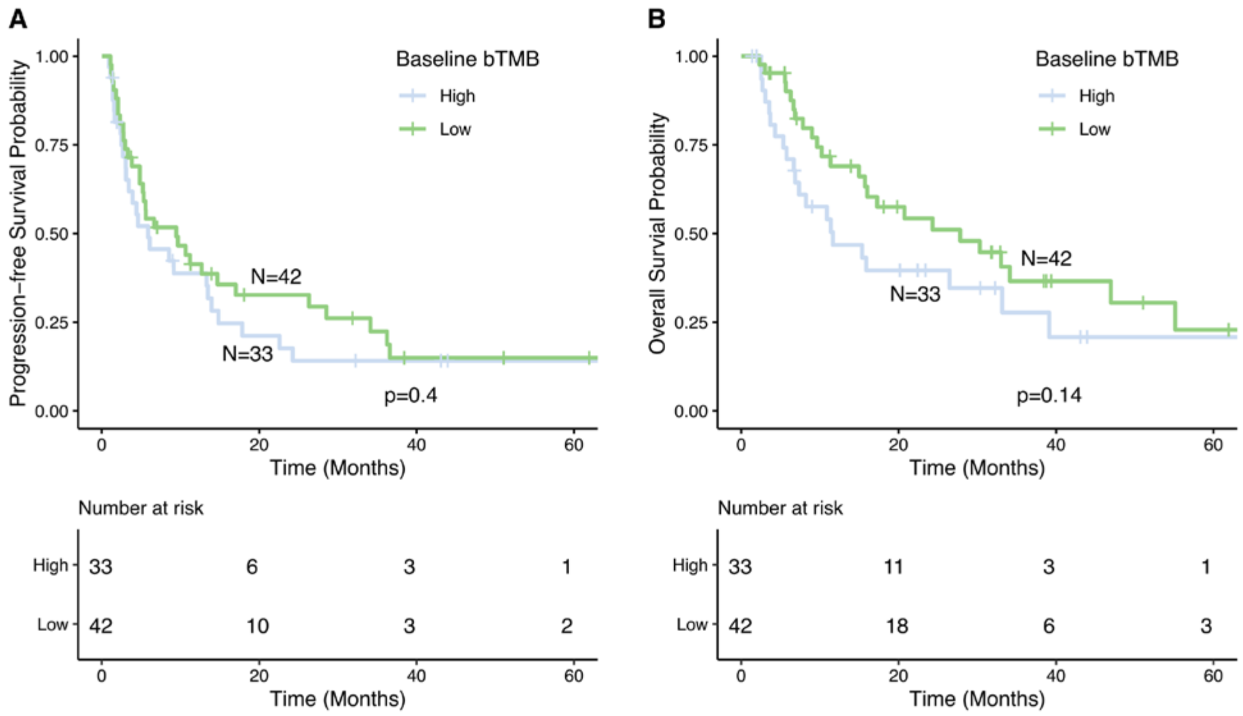


**Figure S6. Association of baseline bTMB, dichotomized at the median, with survival.** Kaplan Meier survival analyses did not reveal a significant association between baseline bTMB and progression-free (A, median 5.9 vs 9.5 months, HR= 0.8 [95% CI 0.48-1.35], logrank P=0.40) or overall (B, median 11.6 vs 27.8 months, HR=0.64 [95% CI 0.36-1.16], logrank P=0.14) survival. Patients were classified as bTMB high and low using the cohort median as the threshold.

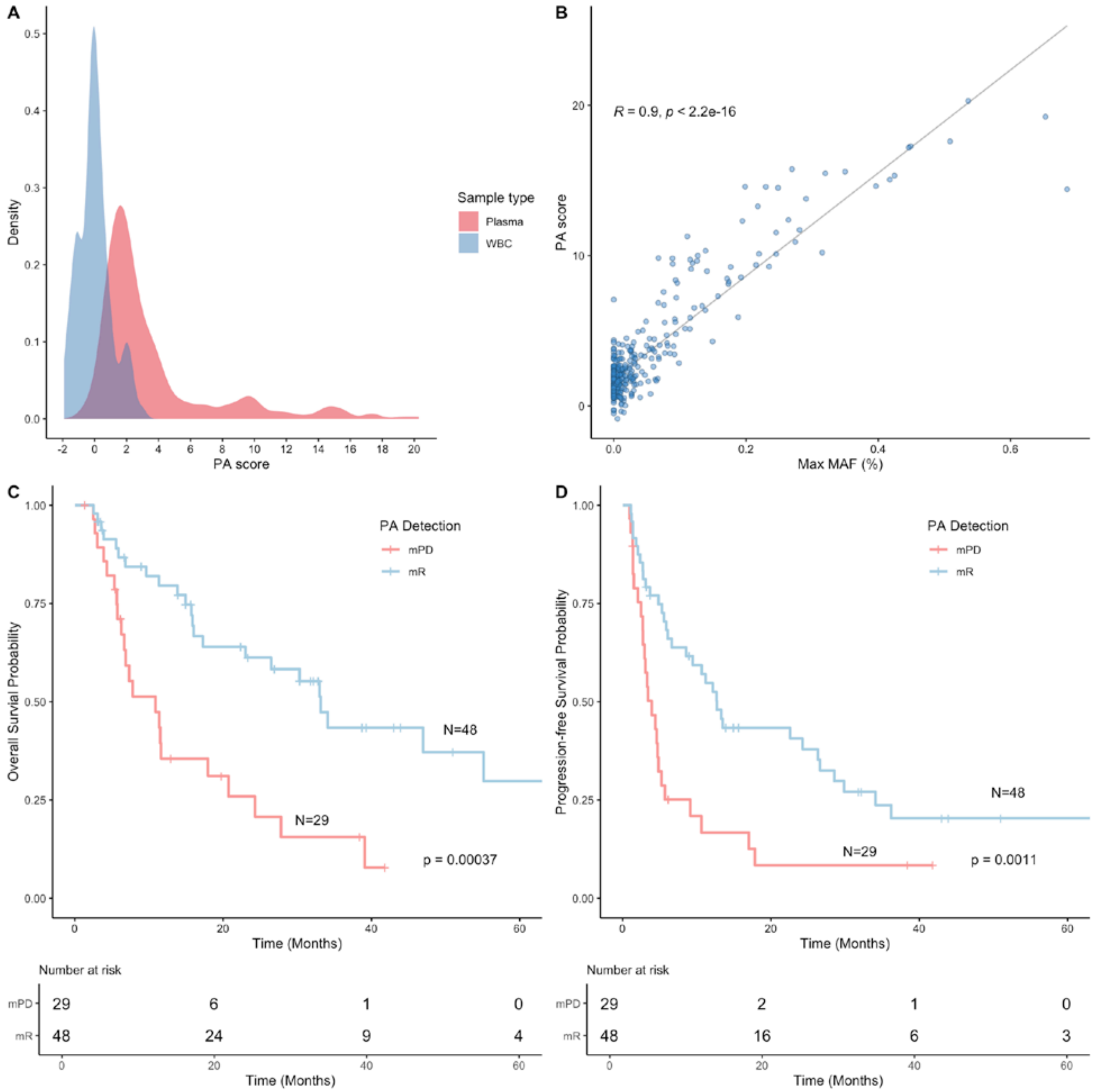


**Figure S7. Assessment of plasma aneuploidy (PA) scores.** (**A**) The distribution of PA scores across plasma and matched white blood cell (WBC) samples analyzed for the study is shown. PA scores from WBC DNA samples ranged from -1.91 to 2.97; therefore, a PA threshold of 2.97 was selected to differentiate between normal ploidy and cancer aneuploidy in plasma (PA scores above 2.97 were indicative of plasma cancer aneuploidy). (**B**) A strong correlation was observed between the maximum mutant allele fraction (maxMAF) of tumor-derived sequence alterations identified across serial plasma samples and segmented plasma aneuploidy scores (Pearson correlation coefficient 0.9058). (**C-D**) Structural responses classified based on landmark assessment of PA scores within 3 -9 weeks of treatment initiation were significantly associated with (C) progression-free survival (PFS) and (**D**) overall survival (OS). Patients with detectable aneuploidy within the landmark window timepoints (PA molecular progression) had a shorter OS (log-rank p = 0.00037) compared to patients with undetectable aneuploidy within the 3–9 week interval (PA molecular response).

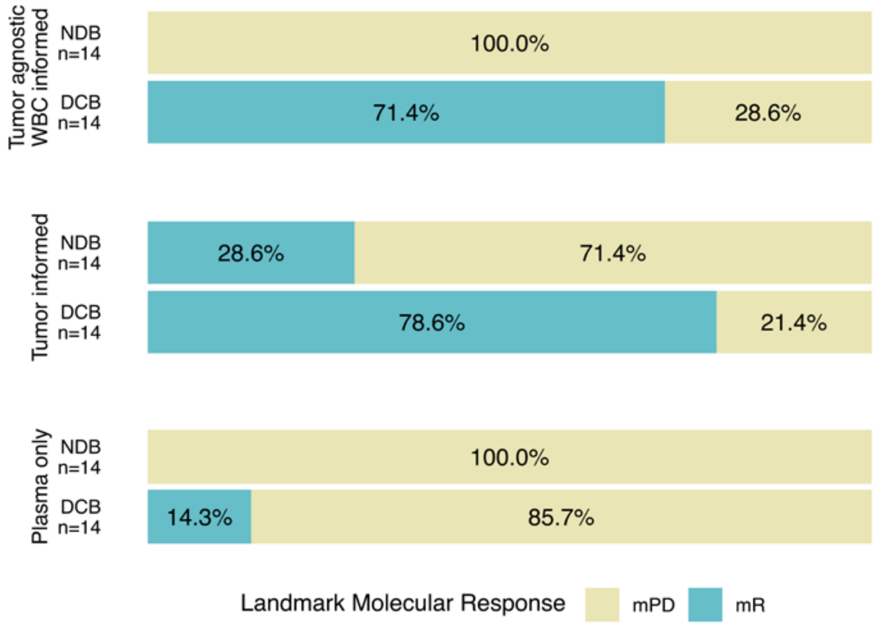


**Fig. S8. Association of landmark molecular response with clinical benefit.** In the subset of patients with available tumor tissue (ν=28), we observed high specificity (100%) for both tumor agnostic WBC informed and Plasma only approaches compared to the tumor informed approach (71.4%). On the other hand, the plasma only approach had a low sensitivity in prediction of clinical benefit (14.3%), while the tumor agnostic WBC informed approach had a sensitivity (71.4%) closer to that of the tumor-informed approach (78.6%).

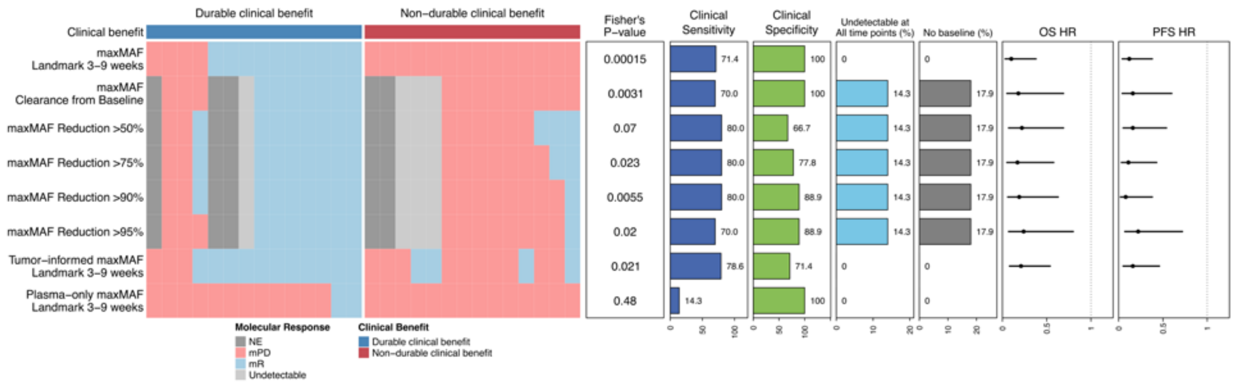


**Fig. S9. Clinical sensitivity of landmark molecular response in comparison to two-timepoint ctDNA response approaches in the subset of patients with tumor tissue.** The heatmap on the right indicates predictions of molecular response for each patient using several approaches. For one-timepoint landmark interval assessment, three methodologies including tumor-agnostic WBC-informed (maxMAF), tumor-informed (Tumor-informed maxMAF), and plasma-only (Plasma-only maxMAF) approaches are assessed. For a two-time-point assessment, clearance or various levels of ctDNA reduction from baseline to the landmark interval are considered. For each approach, the association between clinical benefit and molecular response is evaluated by Fisher's exact test and the p-values are reported, alongside clinical sensitivity and specificity. Clinical sensitivity is defined as the fraction of patients with durable clinical benefit who have a concordant molecular response assignment, and clinical specificity is defined as the fraction of patients with non-durable clinical benefit who have a concordant molecular progressive disease assignment. For the two-timepoint approaches, the fraction of unevaluable patients due to undetectable ctDNA at all timepoints or unavailable baseline sample is reported. Hazard ratios and 95% confidence intervals report the association of each molecular response metric with overall and progression-free survival. Of the subset of 28 patients with available tumor samples shown, 9 patients could not be evaluated using two-timepoint approaches due to baseline unavailability (n=5) or undetectable ctDNA across all time points (n=4). Landmark assessment of molecular response (using tumor-agnostic wbc-informed, tumor-informed, or plasma-only approaches) enabled evaluation of all patients. Landmark tumor-agnostic wbc-informed (denoted as maxMAF landmark 3-9 weeks in the heatmap) yielded a clinical sensitivity similar to that of the tumor-informed approach (71.4% vs 78.6%, respectively) while achieving a significantly higher clinical specificity (100% vs 71.4%, respectively). Conversely, the landmark plasma-only approach had the lowest estimate of clinical sensitivity (14.3%) but was highly specific (100%). With the exception of the landmark plasma-only approach (Fisher's exact p=0.48), all evaluated molecular response definitions were significantly associated with clinical benefit, with the strongest association observed for landmark molecular response by the tumor-agnostic WBC-informed approach (Fisher's exact p=1.5e-4). Similarly, this approach had the strongest association with progression-free (HR=0.12 [95% CI 0.04-0.38], logrank p=4.2e-05) and overall survival (HR=0.10 [95% CI 0.03-0.38], logrank p=8.5e-05), while other definitions were also significantly associated with progression-free and overall survival.

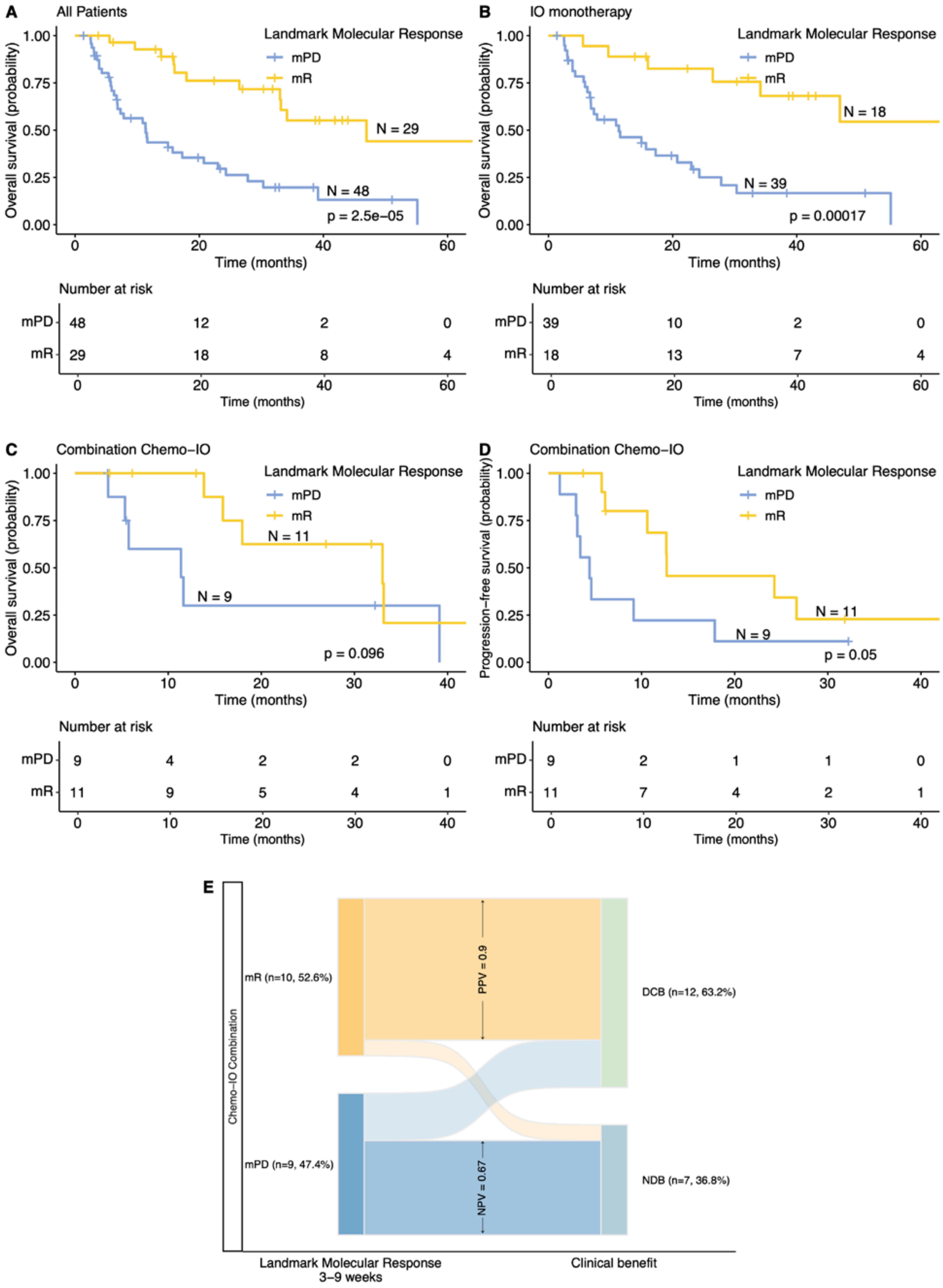
A
All Patients
Landmark Molecular Response
mPD
mR
N = 29
N = 48
p = 2.5e−05
Overall survival (probability)
1.00
0.75
0.50
0.25
0.00
0
20
40
60
Time (months)
Number at risk
mPD 48 12 2 0
mR 29 18 8 4
B
IO monotherapy
N = 18
N = 39
p = 0.00017
Number at risk
mPD 39 10 2 0
mR 18 13 7 4
C
Combination Chemo−IO
N = 11
N = 9
p = 0.096
Number at risk
mPD 9 4 2 2 0
mR 11 9 5 4 1
D
Combination Chemo−IO
Progression−free survival (probability)
N = 11
N = 9
p = 0.05
Number at risk
mPD 9 2 1 1 0
mR 11 7 4 2 1
E
Chemo−IO Combination
mR (n=10, 52.6%)
mPD (n=9, 47.4%)
PPV = 0.9
NPV = 0.67
DCB (n=12, 63.2%)
NDB (n=7, 36.8%)
Landmark Molecular Response
3−9 weeks
Clinical benefit

**Figure S10. Association between landmark ctDNA molecular response and survival outcomes across treatment groups.** Landmark molecular response was associated with longer overall survival across the entire cohort (**A**, median survival of 46.9 vs 11.4 months, logrank p < 0.0001), as well as patients treated with single-agent immunotherapy (**B**, median survival of NR vs 11.3 months, logrank p=0.0002). For patients treated with chemo-immunotherapy combination regimens, the association between landmark molecular response and overall survival (**C)** is shown alongside progression-free survival (**D**). The concordance between molecular response and durable clinical benefit (**E**, p=0.02, Fisher's exact test).
PFS <6 months is annotated as non-durable clinical benefit; NDB.

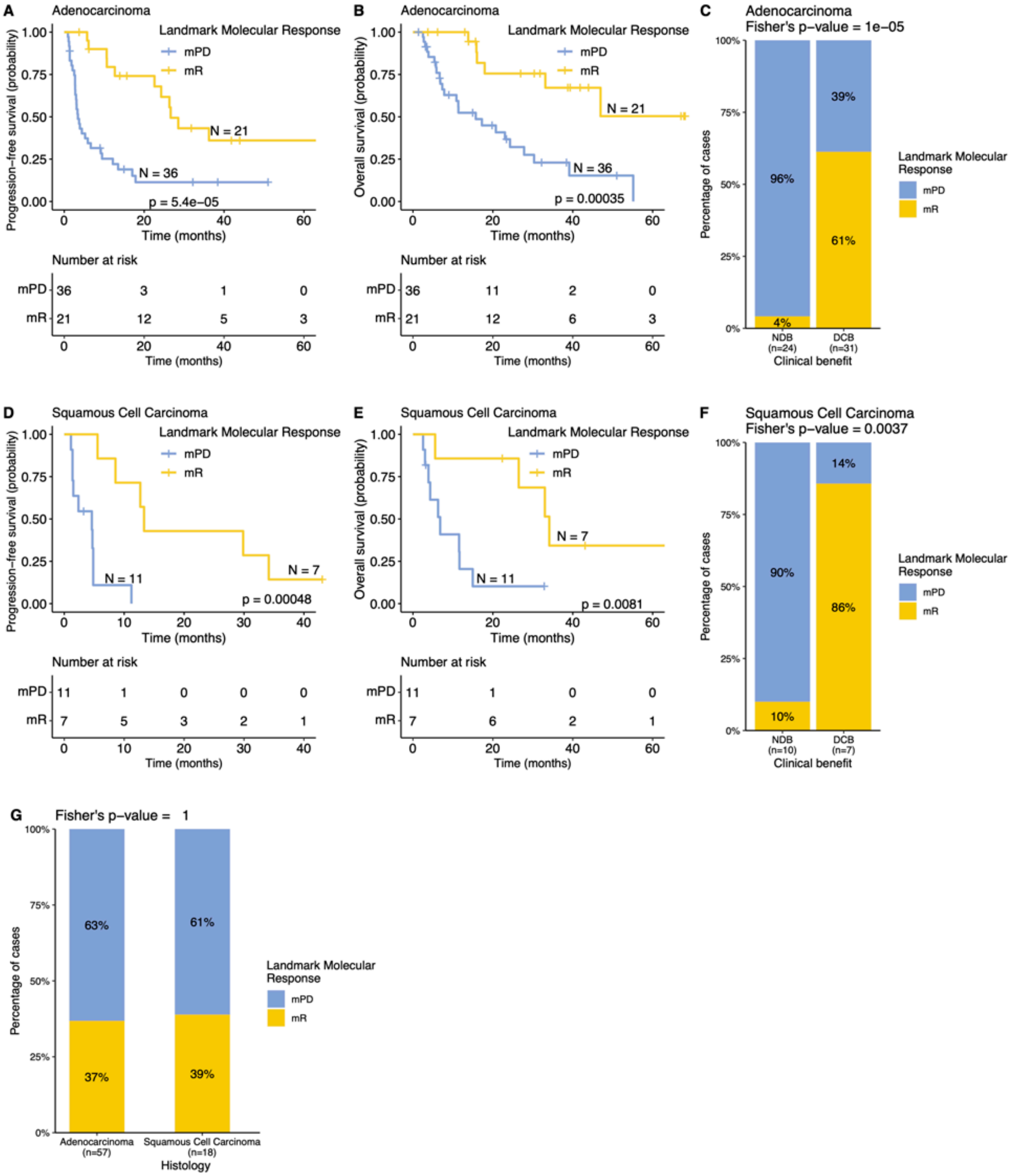


**Figure S11. Association between molecular responses and survival outcomes according to histology.** In patients with adenocarcinoma, molecular responses were significantly associated with progression-free (**A**, median 26.63 vs 3.41 months, logrank p = 5.4e-05) and overall survival (**B**, median NR vs 15.72 months, logrank p < 0.001). ctDNA molecular response predicted durable clinical benefit (PFS ≥6 months) in adenocarcinomas (**C**). Similar associations were observed in patients with squamous subtype tumors (**D**, median progression-free survival of 13.32 vs 4.6 months, logrank p <0.001; **E**, median overall survival of 34.13 vs 6.81 months, logrank p <0.01). ctDNA molecular response predicted durable clinical benefit (PFS

≥6 months) in squamous cell carcinomas (**F**). There was no difference in the molecular response rate between the two histologies (**G**).

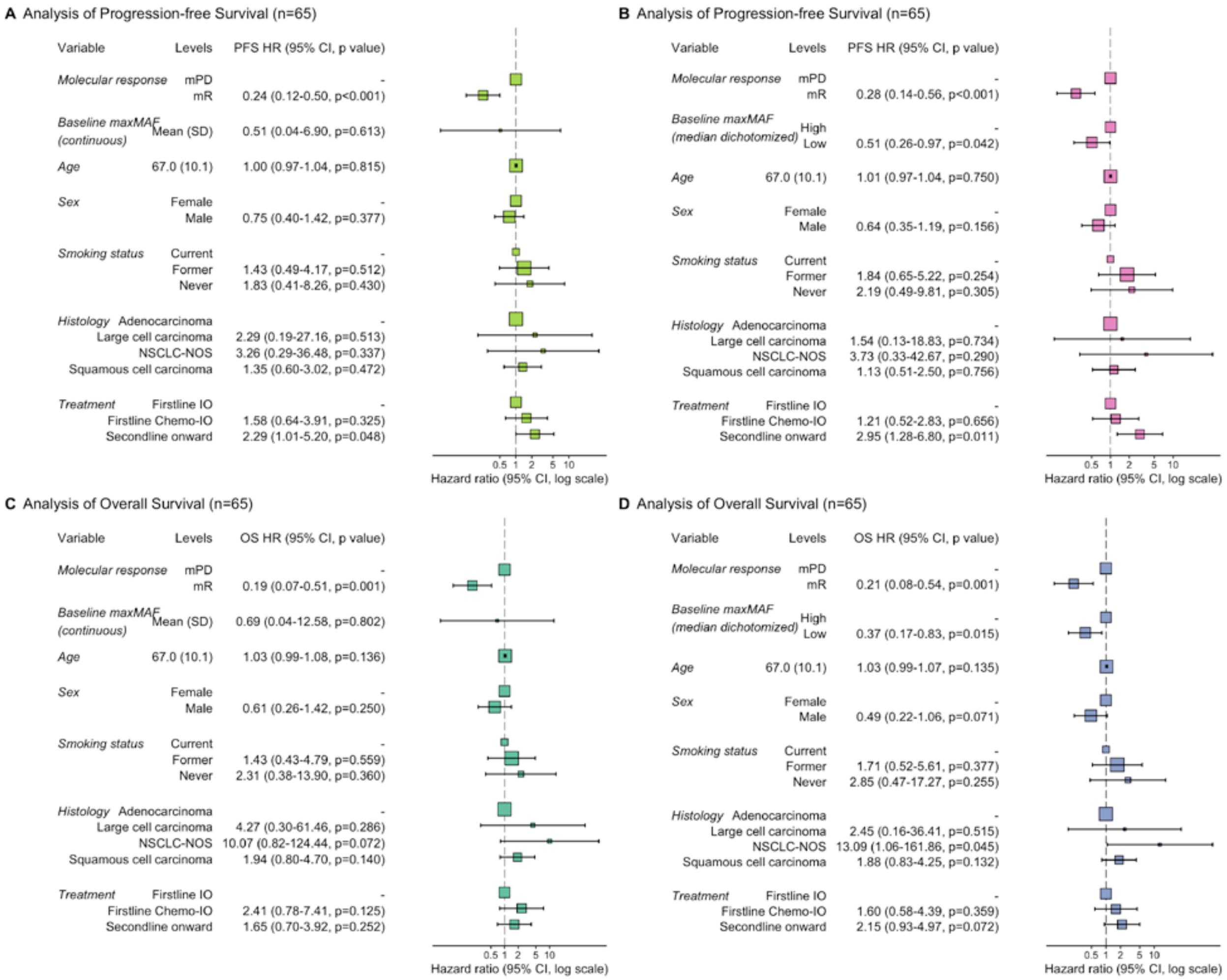


**Figure S12. Independent prediction of progression-free and overall survival for landmark molecular response, adjusting for baseline circulating tumor burden.** We considered two additional approaches to including baseline maxMAF in the multivariate Cox proportional hazards model, either as a continuous variable (**A, C**) or by dichotomizing patients using the cohort median into high- and low-baseline ctDNA groups (**B, D**). In the multivariate model, no significant association was found between baseline maxMAF (treated as a continuous variable) and progression-free (**A**, HR=0.51 [95% CI 0.04-6.90], p=0.61) or overall survival (**C**, HR=0.69 [95% CI 0.04-12.58], p=0.80), while landmark molecular response was a significant predictor (**A**, HR=0.24 [95% CI 0.12-0.50], p<0.001; C, HR=0.19 [95% CI 0.07-0.51], p=0.001). Multivariate models including a dichotomized baseline maxMAF covariate (using the cohort median) found a significant association between progression-free (**B**) and overall (**D**) survival with both landmark molecular response and baseline maxMAF, but the association was stronger for landmark molecular response (PFS, molecular response HR=0.28 [95% CI 0.14-0.56], p<0.001, baseline maxMAF HR=0.51 [95% CI 0.26-0.97], p=0.042; OS, molecular response HR=0.21 [95% CI 0.08-0.52], p=0.001, baseline maxMAF HR=0.37 [95% CI 0.17-0.83], p=0.015). For each clinical or genomic covariate, hazard ratios relative to the reference group, along with 95% confidence intervals and the associated p-value (Wald test), are listed and displayed in the forest plot.
